# How does participation in coal-to-gas policy and availability of natural gas pipelines affect residents’ well-being?

**DOI:** 10.1101/2024.09.08.24313274

**Authors:** Shi Jiankui, HU Lun, Xia Yingge, HU Xiangdong

**Affiliations:** Institute of Agricultural Economics and Development, Chinese Academy of Agricultural Sciences, Beijing, 100081, China; School of Economics and Management, Jiangxi Agricultural University, Nanchang 330044, China; College of Economics and Management, Northwest A&F University, Yangling 712100, China

**Keywords:** coal to gas policy, Pipeline availability, The well-being of the population, New energy transformation effect, Nonlinear relation

## Abstract

With the rapid advancement of industrialization and urbanization, China is facing an increasingly serious challenge of air pollution. The dramatic deterioration of air quality not only compromises the quality of daily life, but also poses a serious threat to public health. In order to deal with the urgent environmental problems, the Chinese government actively seeks treatment methods, among which the coal-to-gas policy, with its advantages of clean and low-carbon, has become a key part of the environmental governance strategy. However, in the initial implementation of the coal-to-gas policy, problems such as shortage of gas sources and pipeline access hindered the promotion of the policy. To provide an in-depth analysis of the actual impact of the coal-to-gas policy on residents’ well-being, a comprehensive analysis was conducted based on data from the China General Social Survey. The study found that the implementation of the coal-to-gas policy not only significantly improved residents’ subjective well-being and made the environment more livable, but also significantly improved residents’ physical health and reduced health problems caused by air pollution. This positive effect is particularly pronounced among young people, women and residents of northern regions, who have benefited more from the clean-air benefits of the coal-to-gas policy. In addition, the study found that the availability of natural gas pipelines has a non-linear effect on residents’ well-being. Moderate pipeline coverage can significantly improve the quality of life of residents and provide more convenient and reliable clean energy. Therefore, under the premise of ensuring stable supply, it is necessary to seek the optimal pipeline layout scheme. Finally, the coal-to-gas policy has boosted regional economic vitality and residents’ well-being by promoting industrial restructuring and reducing pollution emissions.

## 1 Introduction

Over the past few decades, coal energy has generated significant wealth for the global economy, but it has also created new socio-economic problems at the regional and local levels (Gao, 2023). The microscopic matter emitted by coal burning can easily lead to haze. In the Beijing-Tianjin-Hebei region of northern China, the contribution of PM2.5 and SO2 to residential coal burning is as high as 23.1% and 42.6%, respectively (Li et al., 2018), which significantly aggravates air pollution. In addition, the solid combustion process will produce a large number of harmful substances, especially the emission of sulfur, arsenic, fluorine and other elements (Zhao et al., 2018), resulting in acute respiratory infection, COPD, asthma, lung cancer and other diseases (Jin Y et al., 2006). Natural gas is also a fossil energy source. When the corresponding calorific value is generated, the CO2 emissions from burning natural gas only account for 59% of the CO2 emissions from burning coal (Zhou et al., 2021). According to statistics, from 2016 to 2019, the world reduced about 400 million tons of CO2 emissions due to "coal to gas", equivalent to 25% of the total increase in carbon emissions during the same period. The mitigation effect of China’s coal to gas or electricity policy (CNGE) on air quality is estimated to be 3.4μg/m3 PM2.5 and 4.3 μg/m3 SO2 (Weng et al., 2021). The Coal Elimination Initiative is a global convention that limits the environmental and health benefits of coal over the direct policy costs of energy austerity (Rauner et al., 2020).

The well-being of people’s livelihood reflects the aspiration of all mankind for a better life and the ultimate goal pursued by the whole society. His research on people’s well-being in the field of economics can be traced back to the Easterlin period, when he used happiness data to study the relationship between personal happiness and income, and proposed the famous Easterlin Paradox (Easterlin, 2001). As global residents have increasingly high requirements for a sense of gain, security and happiness in life, for developing countries such as China, at this stage, the level of wealth and income is much higher than that of other developing countries (Deaton, 2003; Graham et al., 2017; Yu et al., 2017; Boyce et al., 2010), optimizing social policies (Bonasia et al., 2022), focusing on physical and mental health (Graham et al., 2017) and other non-economic factors are more conducive to improving residents’ well-being. Residents’ self-rated happiness data can also be used to investigate the evaluation effects of public policies (Tella et al., 2006). Air quality has become a key factor affecting residents’ happy life (Zhang X et al., 2017a; Song Y et al., 2019). A large number of studies at home and abroad have shown that air pollution significantly reduces life satisfaction and subjective well-being (Ferreira et al., 2013; Levinson, 2012; Rehdanz et al.), poor air quality can affect hedonic well-being and increase the incidence of depression (Zhang X et al., 2017b).

In terms of energy heating, some countries have basically realized the gradual transition from coal energy to clean energy such as natural gas, in order to deal with the environment and climate change (Song L et al., 2020), and improve the health of residents (Wang Z, 2019) and the level of happiness. In the late 1980s, the South Korean government began to promote alternative energy sources such as kerosene and natural gas. In the late 1990s, natural gas pipelines were laid in most areas of South Korea, and a series of policies and regulations related to environmental protection were promulgated. As the world’s second largest producer of natural gas, the United States has achieved full ventilation in 48 states in 1966, and a complete gas transmission network provides convenience for residents to use natural gas (China Energy Statistical Yearbook). The "14th Five-Year Plan" of modern energy system Planning pointed out that it is necessary to "improve the level of low-carbon electrification of terminal energy use", which covers the field of terminal energy use such as residential life. With reference to Western countries, China has implemented the pilot and promotion policy of "coal to gas", aiming to reduce the emission of environmental pollutants by adjusting the energy structure, which is embodied in replacing some coal with clean energy such as natural gas for residential heating [R/OL].

Although natural gas can provide heat energy and improve the environment in a higher calorific value, low-carbon environmental protection way, and thus help improve residents’ happiness, this alternative way faces some difficulties in the actual implementation of China. The transformation from coal to gas is inseparable from the cost of facilities in the early stage and the cost of gas in the later stage. From the perspective of interregional development, regions with high financial pressure and low income level will face some obstacles in the process of clean energy transition (Zhang Y et al., 2020). Although infrastructure such as natural gas pipelines will be funded by the government, and local residents may be subsidized for gas use, which means that pilot households use high calorific value energy at a low cost, for users, the "coal to gas" project can greatly improve the happiness and health of residents in the pilot area in the early stage. However, considering the long-term natural gas price, future carbon tax, market expansion and other factors (Kemfert et al., 2022), will the government reduce this part of subsidies or completely cancel them? After all, from the perspective of unilateral economic benefits in the Beijing-Tianjin-Hebei region, The cost of natural gas heating is 1.5 to 2 times that of coal heating (Yue et al., 2019), which brings huge financial pressure to the government. At this stage, private gas companies and residential users can hardly bear the high operating costs and gas expenditure (Yue, 2018), which is bound to affect the happy life of residents. Therefore, the impact of the continuous implementation of the "coal to gas" project on the well-being of residents is a topic worthy of in-depth discussion. In terms of energy supply, over-dependence on the energy supply of imported countries and a substantial increase in the demand for central heating in winter may lead to the contradiction between "gas supply shortage" and "excess demand" (Liu et al., 2015; Ruhnau et al., 2023), such problems have indeed occurred in history. The construction of natural gas pipelines and other infrastructure has a certain use cost and threshold, especially in rural areas, the implementation of the "coal to gas" policy is more difficult (Song Y et al., 2019).

The motivation for this paper is rooted in a concern for current global issues of sustainable development and environmental protection, as well as a deep insight into the interconnections between the energy transition and people’s well-being. At a time when global climate change and environmental pressure continue to intensify, promoting the use of clean energy and promoting the optimization and transformation of the energy structure has become an urgent task for the international community and the key to achieving sustainable development. As a major energy consumer in the world, the adjustment and reform of China’s energy policy is of great significance to the global energy pattern and environmental protection. As one of the important strategies in China’s energy structure transformation, the coal-to-gas policy can reduce the use of coal and increase the proportion of natural gas supply and consumption, thus effectively reducing air pollution and improving residents’ living environment. But what impact has this policy had in practice? Does it really improve the well-being of its residents? Is there any difference in the impact of the coal-to-gas policy for different age, gender, region and economic background? These problems are not only the focus of policy makers, but also the topic that needs to be deeply discussed in the field of social science research. Therefore, this paper aims to deeply analyze the specific impacts of the coal-to-gas policy on residents’ subjective well-being and physical health, and carefully discuss the marginal effects, endogeneity, robustness, group heterogeneity and new energy transformation effects of these impacts, so as to provide scientific assessment and suggestions for policy makers, deeply understand the impact mechanism of the coal-to-gas policy, and further optimize the policy design. Promote the optimization and transformation of the energy structure, and realize the comprehensive improvement of residents’ well-being.

The marginal contribution of this paper is that it not only provides a new perspective for understanding the relationship between the coal-to-gas policy and residents’ well-being, but also provides a valuable decision-making reference for policy makers: First, the coal-to-gas policy plays a pivotal role in the process of promoting the energy structure transformation, and its impact on residents’ well-being is far-reaching and complex. In the energy transition, policy design must delicately balance economic, social, and environmental factors. This study reveals a thought-provoking phenomenon: the availability of natural gas pipelines presents a unique "U-shaped" relationship with residents’ well-being. This discovery breaks through the shackles of traditional linear thinking and provides a new thinking dimension for policy makers. As the distance between residents and gas supply points increases, the level of residents’ well-being first shows a downward trend, which warns us of the challenges and obstacles that long-distance supply may bring. However, when the distance reaches a certain level, the level of well-being gradually rises again, which indicates that the compensatory effect of other factors begins to appear under the appropriate distance. This finding requires policy makers to not only pay attention to the full coverage of the macro goals, but also be alert to potential risks and challenges when promoting the coal to gas policy, and ensure that the implementation of the policy can bring environmental benefits and residents’ well-being. Secondly, this paper deeply analyzes the internal mechanism of the coal-to-gas policy affecting residents’ well-being and reveals its multiple benefits. The implementation of the coal to gas policy not only directly improves the living environment and air quality of residents, but also significantly reduces the emission of pollutants generated by coal combustion, thus improving the quality of life of residents; More importantly, it has promoted the transformation of energy consumption structure and industrial upgrading, creating more job opportunities and economic sources for residents. In addition, with the continuous optimization of the energy structure, energy utilization efficiency has been significantly improved, further enhancing the quality of life and well-being of residents. These findings not only enrich our understanding of the effects of coal-to-gas policies, but also provide more comprehensive and in-depth guidance for policy makers. When formulating relevant policies, policymakers need to take into account various factors to ensure that the implementation of policies can maximize their positive impact on the well-being of residents. At the same time, it is also necessary to pay close attention to potential risks and challenges in the process of policy implementation, and take timely and effective measures to deal with them to ensure the smooth promotion and sustainable development of policies. Through continuous optimization of policy design, it is expected that the coal to gas policy will play a greater role in the future and bring more well-being and benefits to residents.

## 2 Literature review and theoretical basis

### 2.1 Participation in the coal-to-gas policy affects residents’ well-being

In the past, scholars studied the impact of the "coal to gas" policy on residents’ well-being from different perspectives. Most of them discussed the environmental benefits of the policy implementation from the perspectives of air quality and environmental governance (Barrington-Leigh et al., 2019; Rauner et al., 2020), and physical health (Wang, 2019; Wu, 2022), and some studies focused on residents’ subjective well-being (Ferreira et al., 2013; Graham et al., 2017; Jin Z et al., 2020). For example, in terms of environmental benefits, household low-carbon energy choice transition contributes to the development of green economy (Niamir et al., 2018). Roger Lueken (Lueken et al., 2016) and others analyzed the replacement of coal by natural gas for power generation in the United States in 2016, and the results showed that the emissions of sulfur dioxide and nitrogen oxides were reduced by more than 90% and more than 60% respectively. Zhang et al(2020) investigated the air quality during China’s clean energy transition period, and found that the concentration of air pollutants such as PM2.5, SO2 and NOx all decreased significantly. The comparison results between the eastern and western regions showed that the emission reduction effect in the western region was more significant than that in the eastern region, except for SO2. Under the Beijing coal reduction plan, although households with different income levels are affected to different degrees in the energy transition, the environmental improvement is effective (Barrington-Leigh et al., 2019). In terms of health benefits, home burning energy substitution helps promote health equity (Fang et al., 2019), can improve self-rated health status of older age groups, and older women benefit more when gender is considered (Wu, 2022). A US study looking at health benefits in terms of economic costs found that switching to natural gas for coal-fired power generation could reduce health-related consumption by $20 billion to $50 billion per year (Lueken et al., 2016). The implementation of the policy ensures people’s livelihood, and the implementation effect is often oriented to the value evaluation at the grassroots level. Compared with the comprehensive effect evaluation of the effectiveness of the "coal to gas" policy before the implementation, the comprehensive effect evaluation has improved in terms of economic cost, environmental benefits, collaborative ecology and health protection, but its core is to improve happiness as the ultimate goal. The satisfaction of public services has an important impact on residents’ happiness (Mu et al., 2023). In the context of air pollution control, the heating mode of "coal to gas" can drive residents to improve their subjective well-being, and the evaluation dimensions of different age groups are not the same. Young people pay attention to life experience and employment security, middle-aged people pay attention to heating subsidies and financial support, and elderly people pay more attention to the actual effect of air governance (Song et al., 2019). Based on the above analysis, this paper proposes the following hypothesis:

Hypothesis 1: Participation in the coal-to-gas policy can significantly improve residents’ physical health and happiness.

### 2.2 The distance of transportation pipeline has U-shaped effect on residents’ happiness

In the fuel transition process, natural gas pipeline networks play an important role in enabling efficient transport to markets and end users (Blanton et al.). In figure 2, (a) and (b) describe the distribution of the number of LNG plants of the magnitude of natural gas pipeline in China, and (c) and (d) show the distribution of natural gas import sites. It can be clearly seen from the figure above that China’s natural gas plants are concentrated in the western region, while most natural gas import sites are located in the eastern coastal region (Lin et al., 2021). Pipeline distance accessibility affects residents’ well-being. On the one hand, from the perspective of countries around the world, regions with natural gas pipelines near coal power plants show higher potential in coal-to-gas projects (Yang Shuting et al., 2022). For example, the regional natural gas pipeline network in the United States is almost 6.5 times that of the interstate highway system (Blanton et al.). It is also a leader in carbon reduction. On the other hand, the construction of natural gas pipeline facilities increases the fixed cost and transportation cost. For example, relying on the Talimu oilfield, Xinjiang has natural energy advantages and competitive price advantages, while the cost of natural gas imported from Xinjiang from Hunan, Jiangxi and other places increases significantly with the increase of transportation distance (Lin et al., 2021). From the above analysis, it can be seen that the key to effectively develop the natural gas pipeline network is to make reasonable use of the location characteristics of the east and west, give play to advantages, and make up for shortcomings. In China, the coal-to-gas project is still in the process of transformation and expansion. Families who live near the natural gas pipeline have better access to participate in the coal-to-gas policy, so they have higher happiness, while families who live far from the natural gas pipeline need to bear higher costs to participate in the coal-to-gas policy, which may lead to lower happiness.

**fig. 1.**
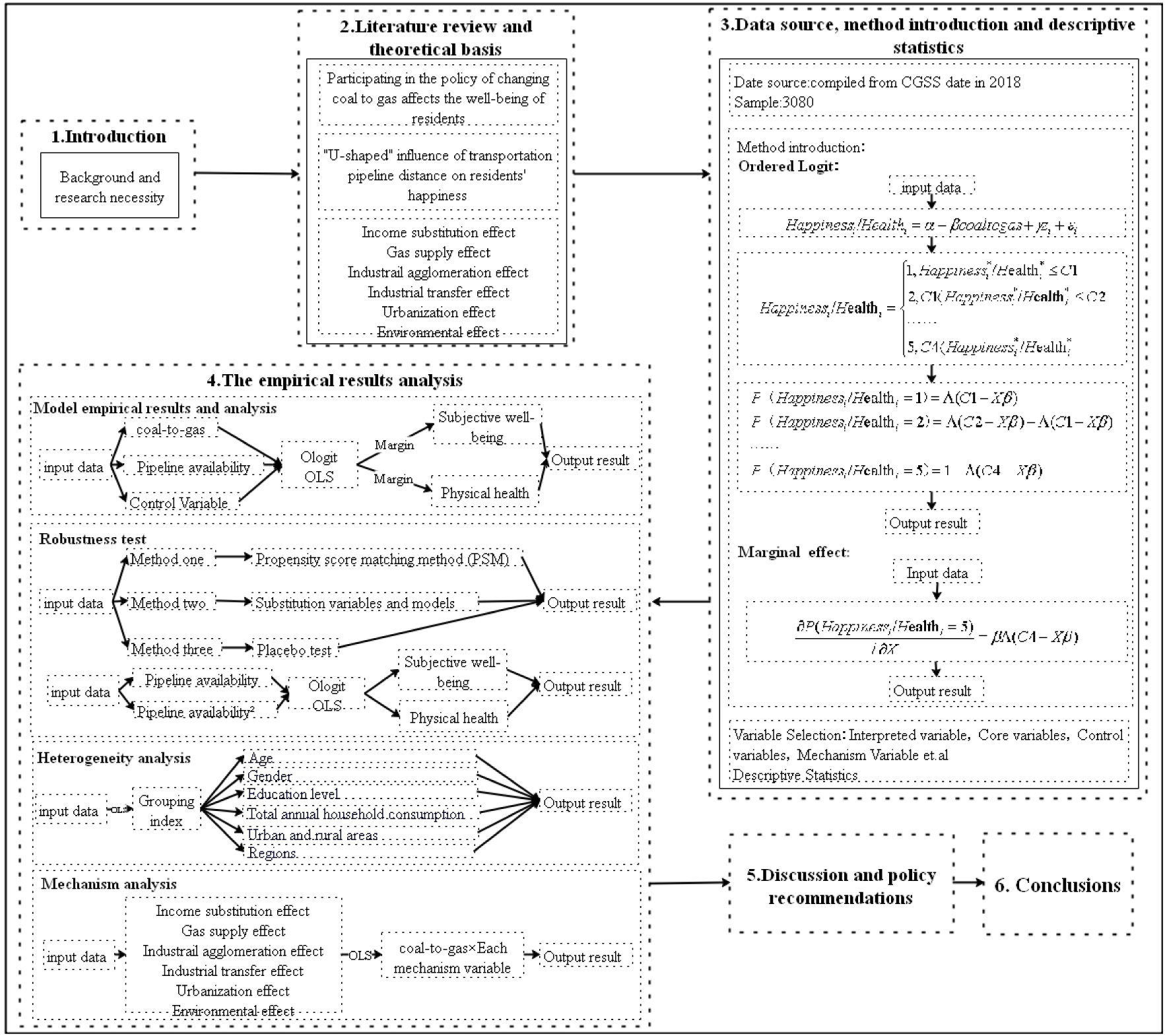
Method flow chart

**fig. 2.**
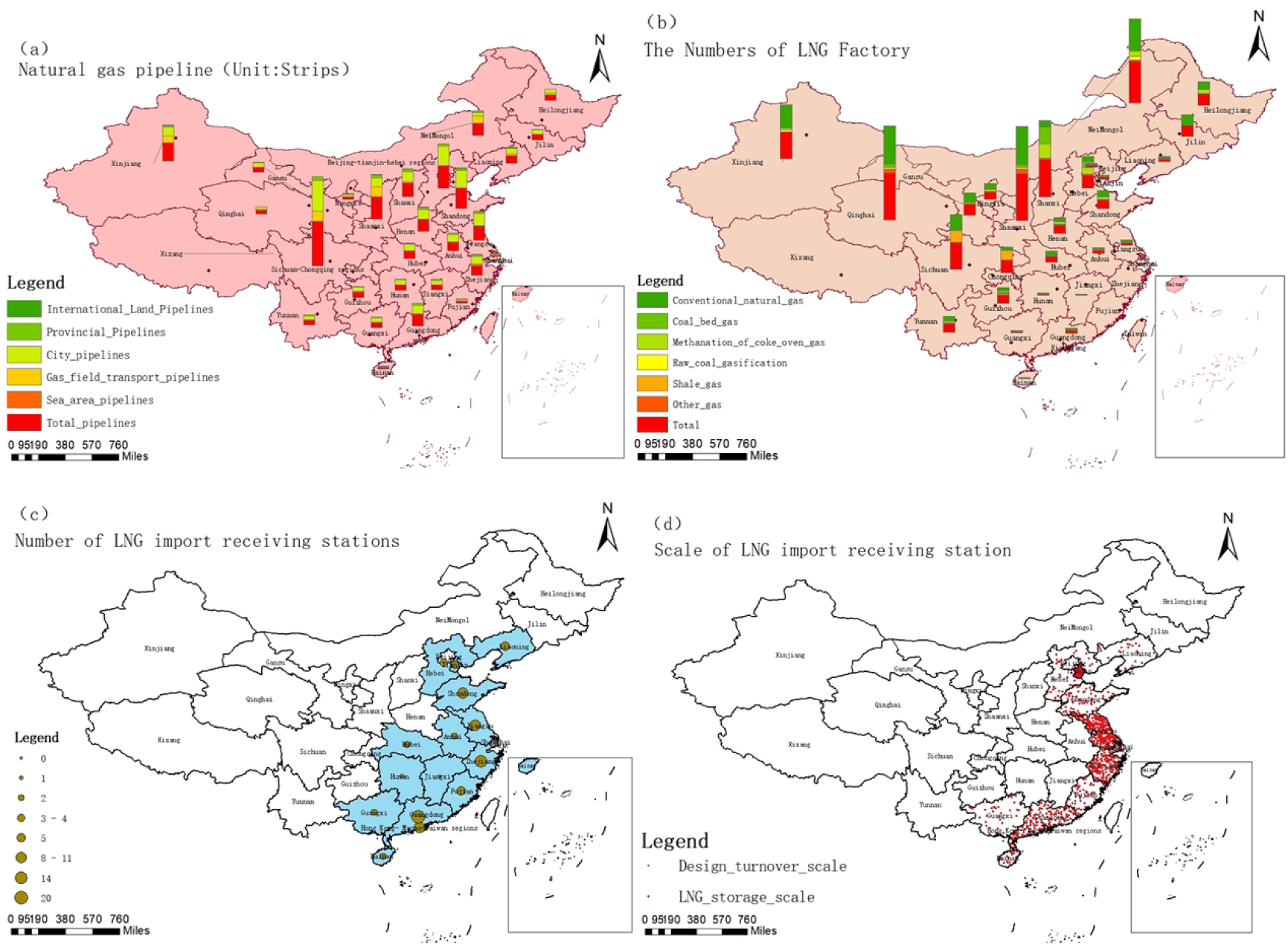
Preliminary analysis on availability of natural gas pipeline. (a)Effective natural gas pipeline projects in 33 provincial administrative regions of China.(b) Effective LNG plant projects in 29 provincial administrative regions of China.(c)Number of effective projects of LNG import receiving stations in 17 provincial administrative regions of China.(d)Scale of LNG import receiving stations in 17 provincial administrative regions of China.(Data source:www.chinagasmap.com)

**fig. 3.**
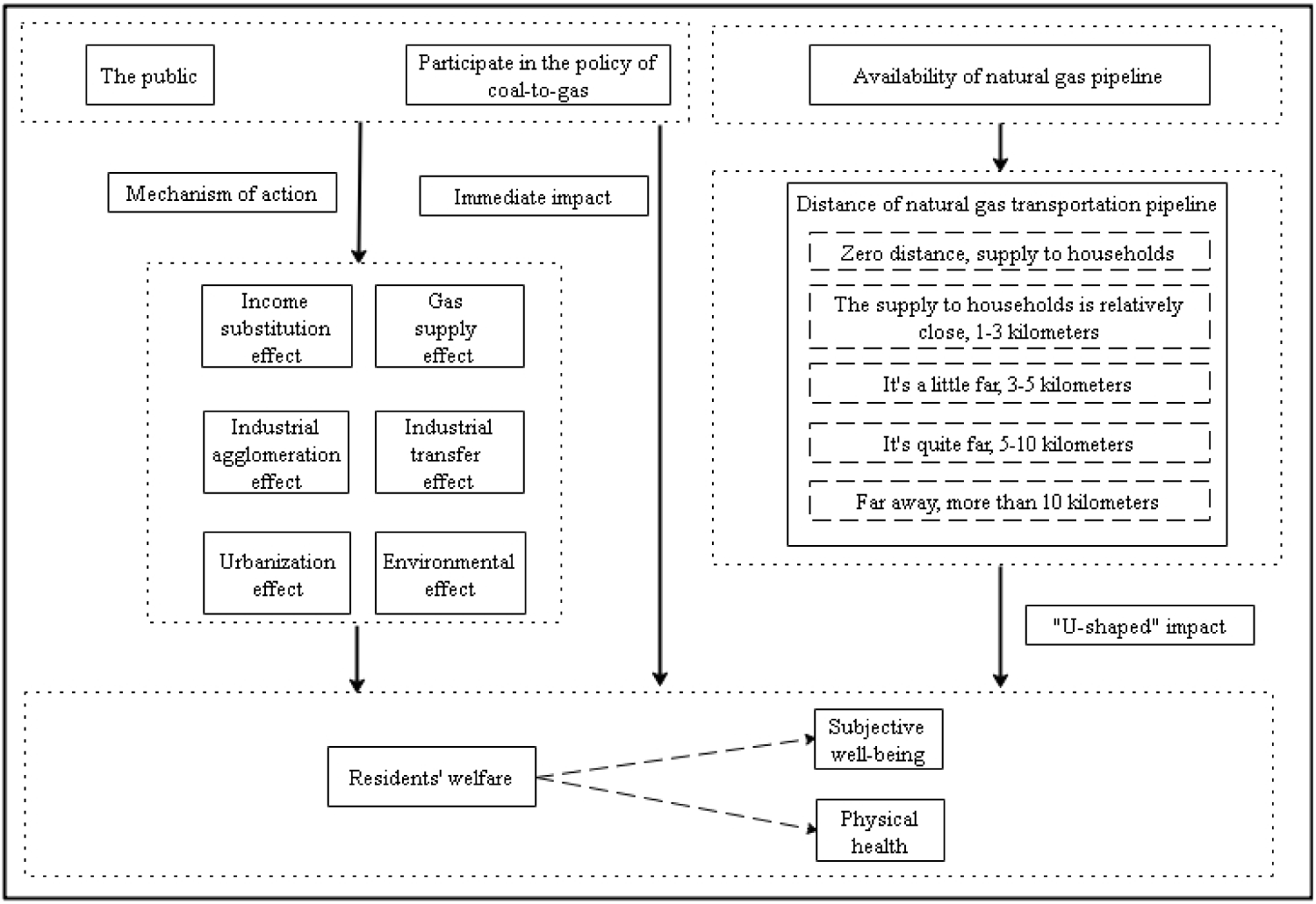
The influence path of participating in the policy of changing coal to gas

However, there is not a simple linear relationship between the distance of natural gas pipeline network and residents’ happiness. As natural gas as a heating fuel occupies most or even all of the market share, natural gas fuel approaches a completely competitive market, and household demand for natural gas and market supply fluctuate steadily for a long time and are at an equilibrium price. At this time, pipeline distance and gas facilities are no longer factors limiting residents’ happiness. In other words, after the influence of pipeline and other factors is excluded, natural gas fuel is used as a heating fuel. Residents’ happiness has been significantly improved, especially families with long-distance pipeline transportation. From the perspective of Europe, which developed earlier, some countries have dispersed gas reserves, although the number of suppliers is limited, and there is no evidence that extreme cold weather has a significant impact on gas prices (Hulshof et al., 2016). It can be seen that with the continuous development of the natural gas market, the ability of all aspects to resist external risks is gradually enhanced, the level of social services is gradually improved, and the happiness of residents is also increased. Based on the above analysis, this paper proposes the following hypothesis:

Hypothesis 2: There is a significant U-shaped relationship between the transportation distance of natural gas pipeline and residents’ happiness, that is, with the increase of transportation distance, residents’ happiness shows a downward trend, and when the transportation distance exceeds a certain point, residents’ happiness shows an upward trend again.

### 2.3 Income substitution effect

Pollution from combustion has negative externalities (Khan M K et al., 2020; Khan S et al., 2021). In order to meet the high requirements of sustainable development, environmental regulations require relevant departments to optimize resource allocation (Wang Y et al., 2016), strengthen the control over polluting industries and clean energy industries, and improve the output efficiency and added value of energy through technological upgrading and equipment transformation (Dong et al., 2018). To achieve the replacement of clean industries to polluting industries, so as to promote households to choose cleaner natural gas energy. Residents are also happier and healthier as a result of the clean energy transition.

From a global perspective, different countries face different policy environments and institutional backgrounds, and the price elasticity of natural gas demand varies between regions and industries (Erias et al., 2022). In China’s residential sector, as a daily heating fuel for households, natural gas belongs to a class of products with inelastic demand prices (Zhang et al., 2018). When natural gas replaces coal as a common heating fuel, natural gas exploitation cost and user price decrease and remain relatively stable. Compared with the early stage of promotion, low natural gas price eases the cost of fuel spent by families and saves part of daily expenditure. Comparatively speaking, per capita disposable income increases, which indicates that families have more choices in other life uses. So it boosts happiness. This paper proposes the following hypotheses:

Hypothesis 3: Participation in the coal to gas policy can improve residents’ happiness by improving regional per capita GDP.

### 2.4 Gas supply effect

Studies have shown that the adoption of administrative powers under government leadership has a positive impact on the demand for natural gas (Cai et al., 2021). Since the demand for natural gas is seasonal, the storage of natural gas can achieve intertemporal arbitrage, thus maintaining a relatively stable price (Hulshof et al., 2016). Residential demand for natural gas fluctuates, and in order to avoid energy shortages, it is necessary to store natural gas energy in summer for use in winter. The implementation of the coal-to-gas policy has significantly increased the demand for natural gas. On the one hand, as the natural gas market continues to expand to grassroots users, the extraction and supply capacity has been significantly enhanced to match the rising demand. On the other hand, natural gas extraction technology is constantly updated and upgraded, which promotes the improvement of social services, especially the level of energy supply services, and thus helps to improve residents’ happiness. This paper proposes the following hypotheses:

Hypothesis 4: Participation in the coal to gas policy will improve residents’ happiness by improving the consumption level and supply capacity of regional air power.

### 2.5 Gas capital intensive industrial agglomeration effect

With the global pursuit of sustainable energy transformation, governments have launched a typical environmental protection policy with coal to gas. (Gao Ying, 2023) states that the governance and perceived authority of local governments will influence issues of energy transition, economic polarization, environmental and social impact. Among them, the gas capital-intensive industry has become the focus of such policies because of its high energy conversion efficiency and low pollution emissions. According to the research of Tao et al., 2024, in some areas of China, the government’s large-scale financial investment in the energy industry leads to the rapid concentration and development of the gas industry, which brings economies of scale, reduces the cost per unit of energy and improves energy efficiency (Liu et al., 2017). In addition, industrial agglomeration provides employment opportunities for local residents and increases job mobility (De Blasio et al., 2005). It also reduces the emission intensity of carbon dioxide (Chen and Chen, 2018), thereby improving residents’ well-being.

Hypothesis 5: Participation in the coal to gas policy is conducive to the rapid concentration and development of gas capital-intensive industries, and the concentration of gas capital-intensive industries has a positive impact on residents’ well-being.

### 2.6 Industrial transfer effect

The coal-to-gas policy led to an increase in the proportion of the tertiary industry and a decline in the degree of industrialization. With the development of the economy and the upgrading of the industrial structure, the proportion of the primary industry may gradually decline, while the secondary industry and the tertiary industry will develop and grow accordingly. Actively promoting industrial restructuring is expected to achieve a "win-win" situation between environmental protection and energy conservation (Tao et al., 2024). As the development of the tertiary industry and the reduction of industrialization also lead to the reduction of air pollution, the coal-to-gas policy can reduce air pollution through industrial structure upgrading and deindustrialization (Yu et al., 2021). In the United States today, coal-based industrial areas remain major population centers, and coal affects the well-being of millions of people in a negative way (Obschonka et al., 2018). Therefore, participation in the coal-to-gas policy can reduce the proportion of coal in industrial industries, reduce the negative impact of coal, and improve residents’ well-being.

Hypothesis 6: Participation in the coal to gas policy has a significant industrial transfer effect, and China’s industrial structure is shifting from the primary industry to the tertiary industry.

### 2.7 Urbanization effect

In most provinces, urbanization trends and gas consumption trends are moving in the same direction, which means there may be a positive correlation. Cai’s research shows that if the urbanization rate increases by 1% in the current year, the natural gas consumption will increase by 0.24% in the following year, which proves that China can effectively promote the natural gas demand by promoting the urbanization process (Cai et al., 2021). At the same time, urbanization and natural gas consumption have a long-term two-way causal relationship. In China, the thermal insulation performance of most rural houses is poor, and most residents use coal and wood for heating, which is quite an obstacle in terms of safety performance and convenience of life. Participation in coal gas transformation can improve residents’ heating security and home environment to a certain extent, and improve the level of urbanization (Bai et al., 2023). Participation in the coal-to-gas policy has a positive impact on the happiness of residents, and the happiness of residents participating in the coal-to-gas policy will be significantly improved.

Hypothesis 7: Participation in the transformation from coal to gas has a positive impact on residents’ happiness by improving the level of urbanization.

### 2.8 Environmental effect

The implementation of the coal to gas policy is closely related to the reduction of air pollution (Zeng et al., 2022), and the emissions of sulfur dioxide and PM2.5 in cities that switch to natural gas generally decrease. Research (Lueken et al., 2016) shows that switching to natural gas in the United States will greatly reduce the emission of standard pollutants, while sulfur dioxide emissions will be reduced by more than 90%. The Chinese government plans to reduce air pollution in northern China by switching from coal to natural gas for winter heating, and the results also confirm that Beijing’s energy transition has effectively reduced air pollution (Fan et al., 2020). (Yu et al., 2021) showed that participation in the coal-to-gas policy resulted in a 31.3% decrease in industrial sulfur dioxide and a 36% decrease in industrial smog. At the same time, after eliminating the spatial diffusion interference through SLM-DID, the coal-to-gas policy could reduce the quality of PM2.5 by 7%. The coal to gas policy has obvious improvement effect on air pollution. Under the influence of switching to natural gas, the emission of standard pollutants in the United States has been reduced and the health of residents has been improved (Lueken et al., 2016). (Fan et al., 2020) also shows that air pollution increases the death rate of asphyxia in winter, and reducing air pollution is conducive to reducing the death rate and improving residents’ well-being.

Hypothesis 8: Participation in coal to gas conversion can improve residents’ happiness by reducing the emission of smoke and dust in the air.

## 3 Data sources, methods and descriptive statistics

### 3.1 Data source

The data in this paper are derived from CGSS data of 2018. Since its launch in 2003, China General Social Survey (CGSS) has gradually established itself as an indispensable tool in the field of social science research. It not only draws the essence of the US General Social Survey (GSS), but also makes in-depth innovation and localization adjustment on the basis of China’s unique national conditions. In particular, the CGSS data set of 2018 provides a detailed record of rich information on Chinese people’s well-being, traditional energy consumption patterns, and the specific impact of policy adjustments (such as coal-to-gas policies) on people’s lives (Wang et al.,2020). As the first comprehensive large-scale social survey project covering the whole country in China, CGSS adopts the scientific multi-layer probability sampling method in sampling design to ensure the breadth and representativeness of samples. The survey uses the advanced multi-layer probability sampling method to carefully select samples across the country. It covers 28 provincial-level administrative units in China except Hong Kong, Macao and Taiwan, Xinjiang, Tibet and Hainan. Through this systematic and comprehensive survey, CGSS aims to accurately reflect the basic conditions of all aspects of Chinese society and provide valuable empirical data for social science research. At present, CGSS data has been widely recognized by the academic community and is regarded as representative data with scientific research value, which provides solid support for in-depth understanding and analysis of structural changes, dynamic mechanisms and the formation of social problems in Chinese society.

### 3.2 Method introduction

Residents’ well-being and physical health are typical discrete ordering data. This paper followed the international general processing method and adopted the Ordered Logit (Ologit) model for regression. The Ologit model treats happiness as a ranking variable, and the estimation model is as follows:

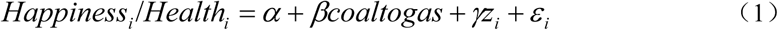

*Happiness_i_* and *Health_i_* represent the happiness of the *i* resident, and *coaltogas_i_* represents the *i* resident’s participation in the transformation from coal to gas. *_Zi_* is the control variable (including age, gender, education, marriage, household registration, total household income, family population size, total household expenditure, understanding degree of coal to gas, etc.).

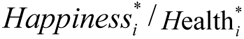 is the latent variable, residents feel very unhappy/very unhealthy when 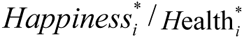 is below the critical value of *C*1 (*Happiness_i_* /*Health_i_* = 1), less happy/less *i*healthy when above *C*1 but below *C*2 (*Happiness_ii_*/*H*ealth*_i_* = 2), and so on, residents feel very happy/very healthy when above *C*4 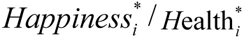. As shown in equation (2) :

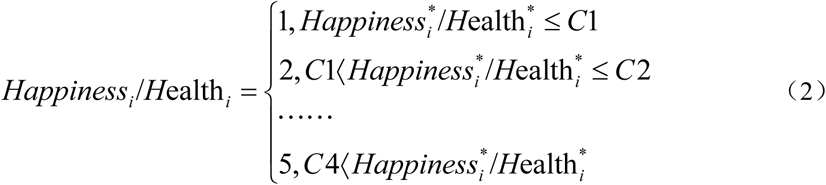

Assuming that *ε_i_* is subject to a logical distribution, *X* represents all explanatory variables, and ∧(·) represents the cumulative distribution function, then *Happiness_i_* /*H*ealth*_i_* can be expressed as:

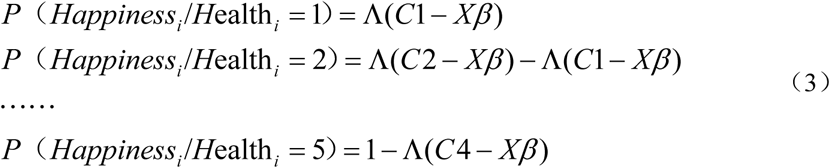

Further, because the coefficient estimated by the Ologit model is not intuitive and can only give limited information in terms of significance and parameters, the marginal effects of each explanatory variable on well-being and physical health reported in our empirical analysis are as follows:

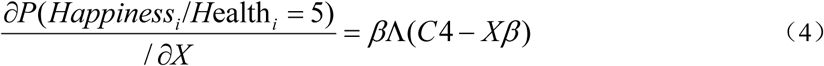

### 3.3 Descriptive statistics

1. The core explained variables of this study are residents’ happiness and physical health, which jointly measure residents’ well-being (Yin et al.,2024). According to the questionnaire, "Overall, do you feel happy with your life?" Combined with the options "very unhappy, relatively unhappy, not happy or unhappy, relatively happy or very happy", the corresponding values are assigned, "very unhappy" is assigned 1, "relatively unhappy" is assigned 2, "not happy or unhappy" is assigned 3, "relatively happy" is assigned 4, and "very happy" is assigned 5. Thus, the ranking features reflecting residents’ happiness are formed. "Overall, do you think you are Physical health?" In combination with the options "very unhealthy, relatively unhealthy, not healthy, not healthy, relatively healthy, very healthy", the corresponding value is assigned, "very unhealthy" is assigned 1, "relatively unhealthy" is assigned 2, "not healthy or unhealthy" is assigned 3, "relatively healthy" is assigned 4, and "very healthy" is assigned 5. Thus, the ranking features reflecting residents’ physical health are formed. The statistics of residents’ happiness and physical health are shown in Table 1 below.

**Table. 1.** Statistics of residents’ happiness and physical health.
2. Core explanatory variables: participation in the coal-to-gas policy and pipeline availability. The questionnaire question is set to "Whether your family has participated in the coal to gas project", with a value of 1 for participation and 0 for non-participation. figure 4 and figure 5 show the spatial distribution of happiness and physical health of residents who participate in the coal to gas transformation and those who do not. The preliminary results show that the happiness and physical health of residents in the southern region are significantly higher than those in the northern region, and the participation in the coal to gas transformation policy will significantly improve the happiness and physical health of residents in the whole country, especially in the northern region. Gansu and Yunnan regions have the highest happiness of life when they participate in the coal to gas transformation. In the case of coal to gas conversion, Qinghai and Yunnan regions have the best health status. Based on the study of Yang et al.(2022), pipeline availability is selected as another core explanatory variable. According to the distance from the natural gas transportation distance "zero distance, supply to households; Supply to the household is relatively close, 1-3 km; It’s a bit far, 3-5 km; Relatively far, 5-10 km; "Far away, more than 10 kilometers" is assigned a value of 1 to 5.
3. Control variables. The main indicators that affect residents’ happiness of life include social development characteristics, such as individual socio-demographic characteristics and macro environmental characteristics of the city. Therefore, it is necessary to introduce control variables into the model. Individual characteristic variables were controlled from education level, gender, age, marriage and household registration (Hu et al,2022). The degree of understanding from coal to gas, the pollution degree of energy products are different, and the environmental protection sacrifices economic benefits (Shi and Hu,2024) to control individual environmental cognitive variables. From the family characteristics variables of total household income, family population size, and total household expenditure control (Wang et al.,2021); From the fourth, it is divided into eastern, central and western (Hu et al.2022) control region dummy variables (taking Northeast China as an example).
4. Mechanism variables. In order to explore the impact mechanism of participation in the coal-to-gas policy on residents’ well-being, income substitution effect (Yu et al.,2021), gas supply effect, gas capital-intensive industry agglomeration effect, industrial transfer effect, urbanization effect, and environmental effect were selected as the mechanism variables to discuss, starting from objective and scientific considerations. All the variables in this paper are summarized in Table 2.

| Happiness of life | Sample size | Frequency (%) |
| --- | --- | --- |
| Very unhappy | 70 | 2.27 |
| Less happy | 333 | 10.81 |
| can't say happy or not | 508 | 16.49 |
| Relatively happy | 1630 | 52.92 |
| Very happy | 539 | 17.5 |
| Physical health | Sample size | Frequency (%) |
| Very unhealthy | 122 | 3.96 |
| Less healthy | 678 | 22.01 |
| normal | 925 | 30.03 |
| Relatively healthy | 892 | 28.96 |
| Very healthy | 463 | 15.03 |

**Table. 2.**
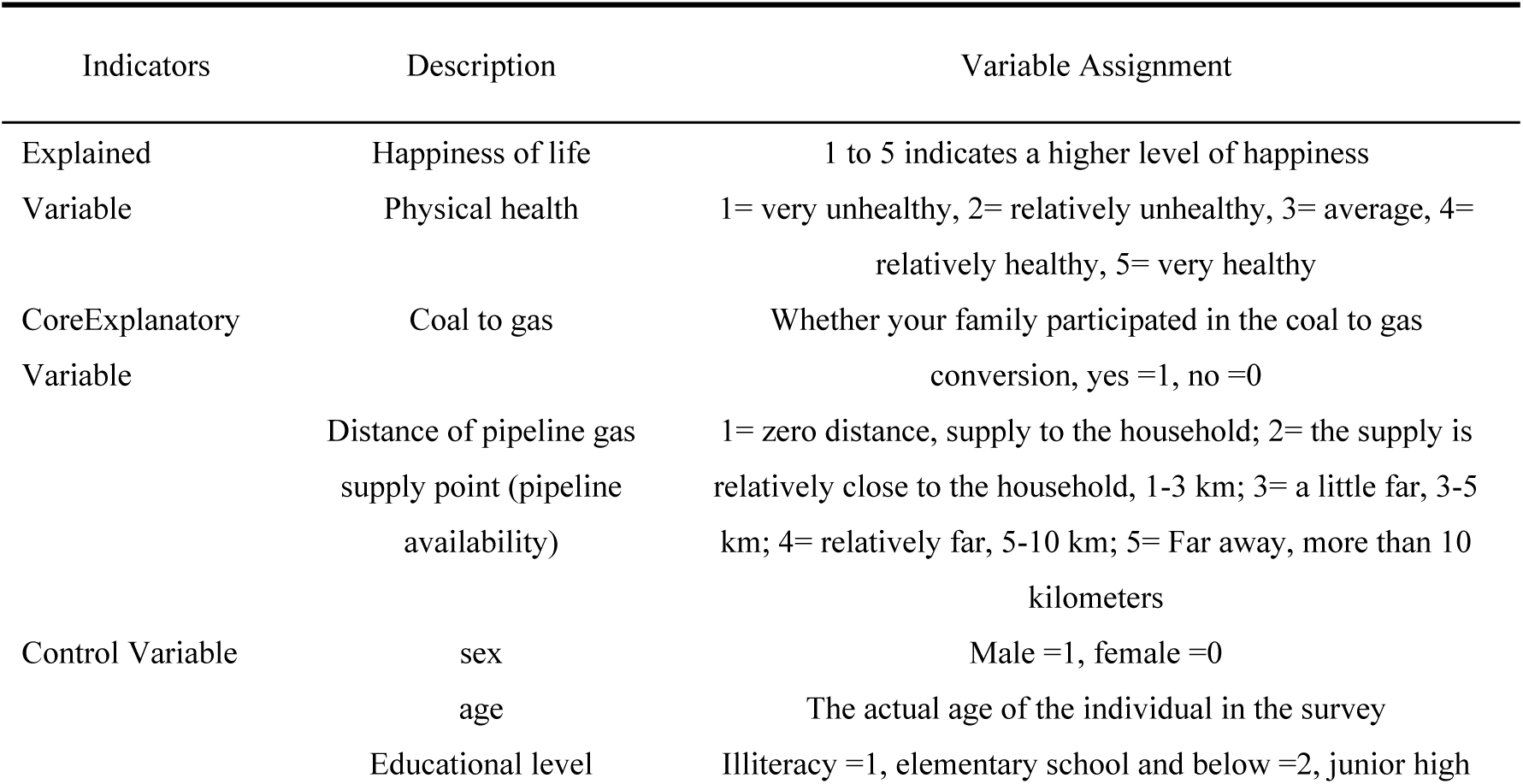
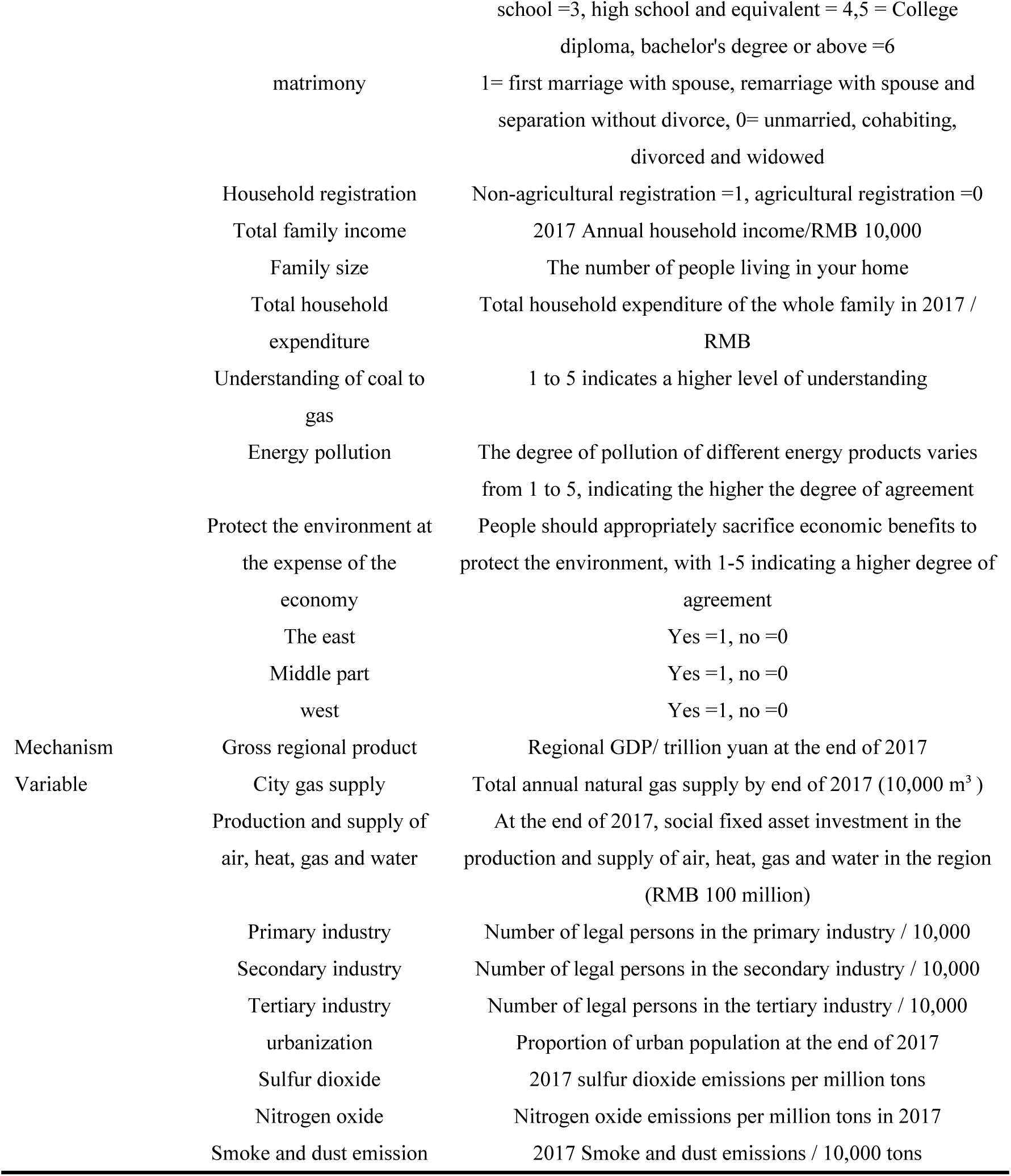
Variable assignment.

As can be seen from Table 3, the average well-being of residents who do not participate in the coal to gas policy is significantly lower than that of residents who participate in the coal to gas policy. The two groups of samples showed significant differences in gender, age, education level, marriage, household registration, total household income, total household expenditure, understanding degree of coal to gas, energy pollution, economic degree of environmental protection sacrifice, eastern, central, western, gross regional product, primary industry, secondary industry, tertiary industry, urbanization, sulfur dioxide and smoke dust emissions. Residents who participate in the coal to gas policy have higher well-being, and most of them are male, relatively healthy, and their family economic status is relatively good.

**fig. 4.**
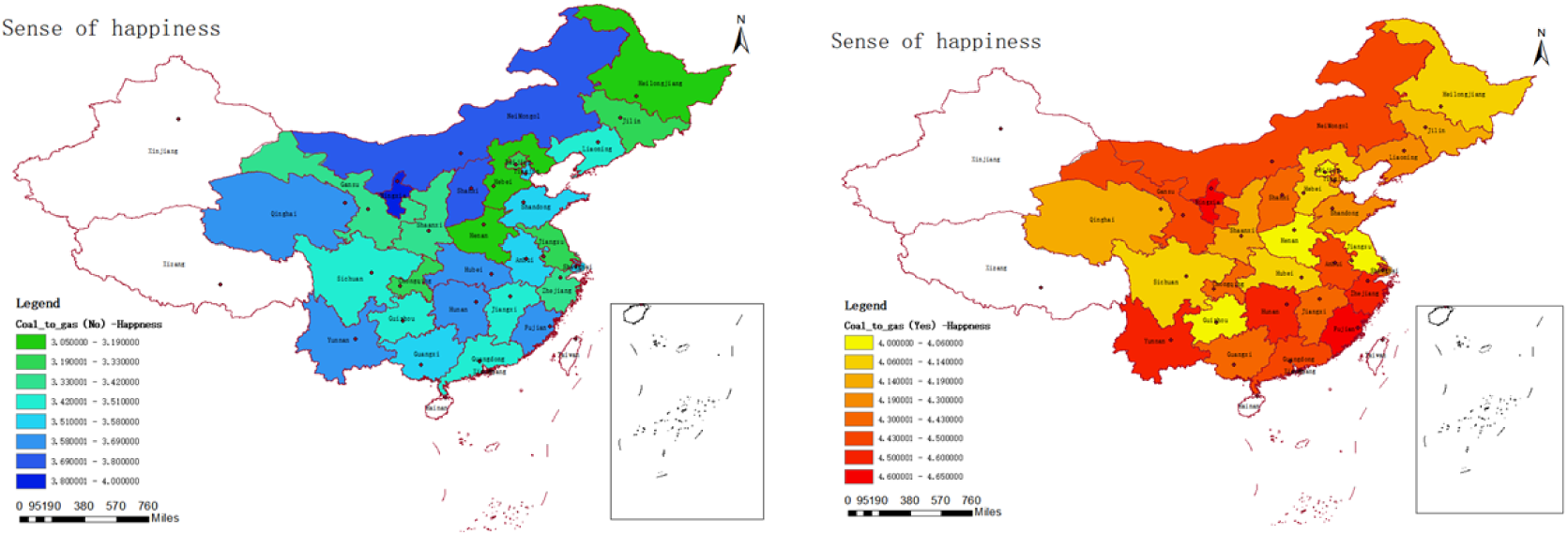
National difference of happiness who participate in the coal-to-gas policy or not

**fig. 5.**
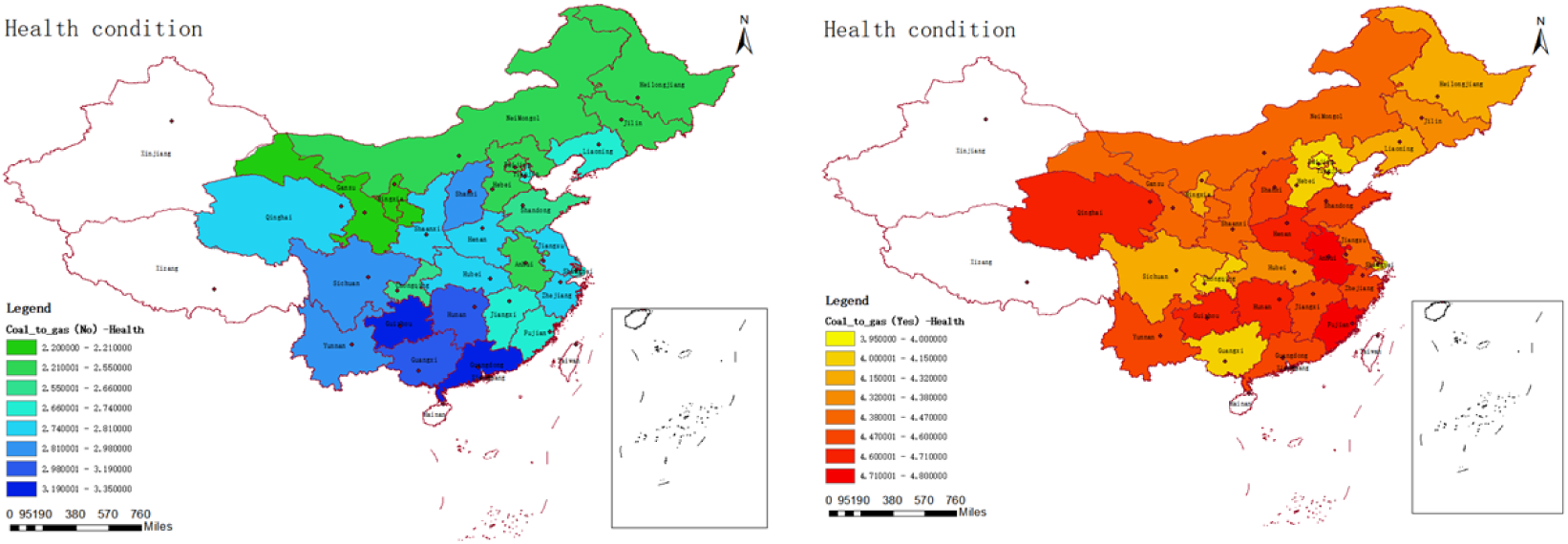
National differences in health of residents who participate in the coal-to-gas policy

**Table. 3.** Difference test.

| Description | Total sample |  | participant |  | Have not participated |  | T-value test |
| --- | --- | --- | --- | --- | --- | --- | --- |
|  | Mean | S.D. | Mean | S.D. | Mean | S.D. |  |
| Happiness of life | 3.726 | 0.95 | 4.229 | 0.638 | 3.464 | 0.98 | -0.765*** |
| Physical health | 3.291 | 1.089 | 4.309 | 0.65 | 2.762 | 0.874 | -1.547*** |
| Distance of pipeline gas supply point<br>(pipeline availability) | 1.494 | 1.015 | 1.47 | 1.057 | 1.517 | 0.974 | 0.047** |
| sex | 0.469 | 0.499 | 0.481 | 0.5 | 0.463 | 0.499 | -0.018*** |
| age | 51.461 | 16.183 | 47.572 | 16.466 | 53.481 | 15.662 | 5.909*** |
| Educational level | 3.055 | 1.253 | 3.484 | 1.169 | 2.831 | 1.238 | -0.653*** |
| matrimony | 0.784 | 0.411 | 0.799 | 0.401 | 0.777 | 0.416 | -0.022*** |
| Household registration | 0.378 | 0.485 | 0.479 | 0.5 | 0.326 | 0.469 | -0.153*** |
| Total family income | 10.518 | 1.784 | 11.198 | 1.116 | 10.165 | 1.955 | -1.033*** |
| Family size | 2.812 | 1.388 | 2.862 | 1.361 | 2.786 | 1.401 | -0.076 |
| Total household expenditure | 10.136 | 1.061 | 10.305 | 0.988 | 10.048 | 1.087 | -0.257*** |
| Understanding of coal to gas | 2.492 | 1.317 | 3.206 | 1.263 | 2.121 | 1.186 | -1.085*** |
| Energy pollution | 3.866 | 0.693 | 4.019 | 0.591 | 3.786 | 0.728 | -0.233*** |
| Protect the environment at the expense of<br>the economy | 3.695 | 0.844 | 3.818 | 0.805 | 3.631 | 0.857 | -0.187*** |
| The east | 0.38 | 0.485 | 0.462 | 0.499 | 0.337 | 0.473 | -0.125*** |
| Middle part | 0.241 | 0.428 | 0.211 | 0.408 | 0.257 | 0.437 | 0.046*** |
| west | 0.235 | 0.424 | 0.13 | 0.337 | 0.29 | 0.454 | 0.16*** |
| Gross regional product | 10.209 | 0.591 | 10.182 | 0.603 | 10.223 | 0.585 | 0.041*** |
| City gas supply | 12.804 | 0.939 | 12.962 | 0.86 | 12.723 | 0.967 | -0.239*** |
| Production and supply of air, heat, gas and<br>water | 6.716 | 0.683 | 6.655 | 0.718 | 6.747 | 0.662 | 0.092 |
| Primary industry | 10.655 | 0.903 | 10.473 | 0.964 | 10.749 | 0.855 | 0.276*** |
| Secondary industry | 11.678 | 0.831 | 11.607 | 0.864 | 11.715 | 0.812 | 0.108*** |
| Tertiary industry | 13.134 | 0.766 | 13.102 | 0.741 | 13.151 | 0.778 | 0.049*** |
| urbanization | 4.118 | 0.193 | 4.157 | 0.198 | 4.098 | 0.187 | -0.059*** |
| Sulfur dioxide | 2.928 | 1.083 | 2.821 | 1.249 | 2.983 | 0.981 | 0.162*** |
| Nitrogen oxide | 3.615 | 0.59 | 3.614 | 0.67 | 3.616 | 0.543 | 0.002 |
| Smoke and dust emission | 2.908 | 0.926 | 2.864 | 1.125 | 2.932 | 0.804 | 0.068*** |

## 4 Empirical result analysis

### 4.1 Participate in the coal-to-gas policy and residents’ well-being

Table 4 shows that whether the Ologit model or OLS model regression is used, the coal-to-gas policy has a significant positive impact on residents’ well-being. In other words, compared with residents who do not participate in the coal-to-gas policy, residents who participate in the coal-to-gas policy have higher well-being, with typical environmental effects. Participating in the coal-to-gas policy can reduce the use of coal, promote the use of clean energy, significantly improve air quality, improve residents’ health and well-being, reduce economic burden, optimize the energy structure, and thus enhance residents’ quality of life and energy security. Thus, the sustainable development of society is promoted, and hypothesis 1 is verified, which is consistent with the research conclusion of Shi and Hu (2024).

**Table. 4.**
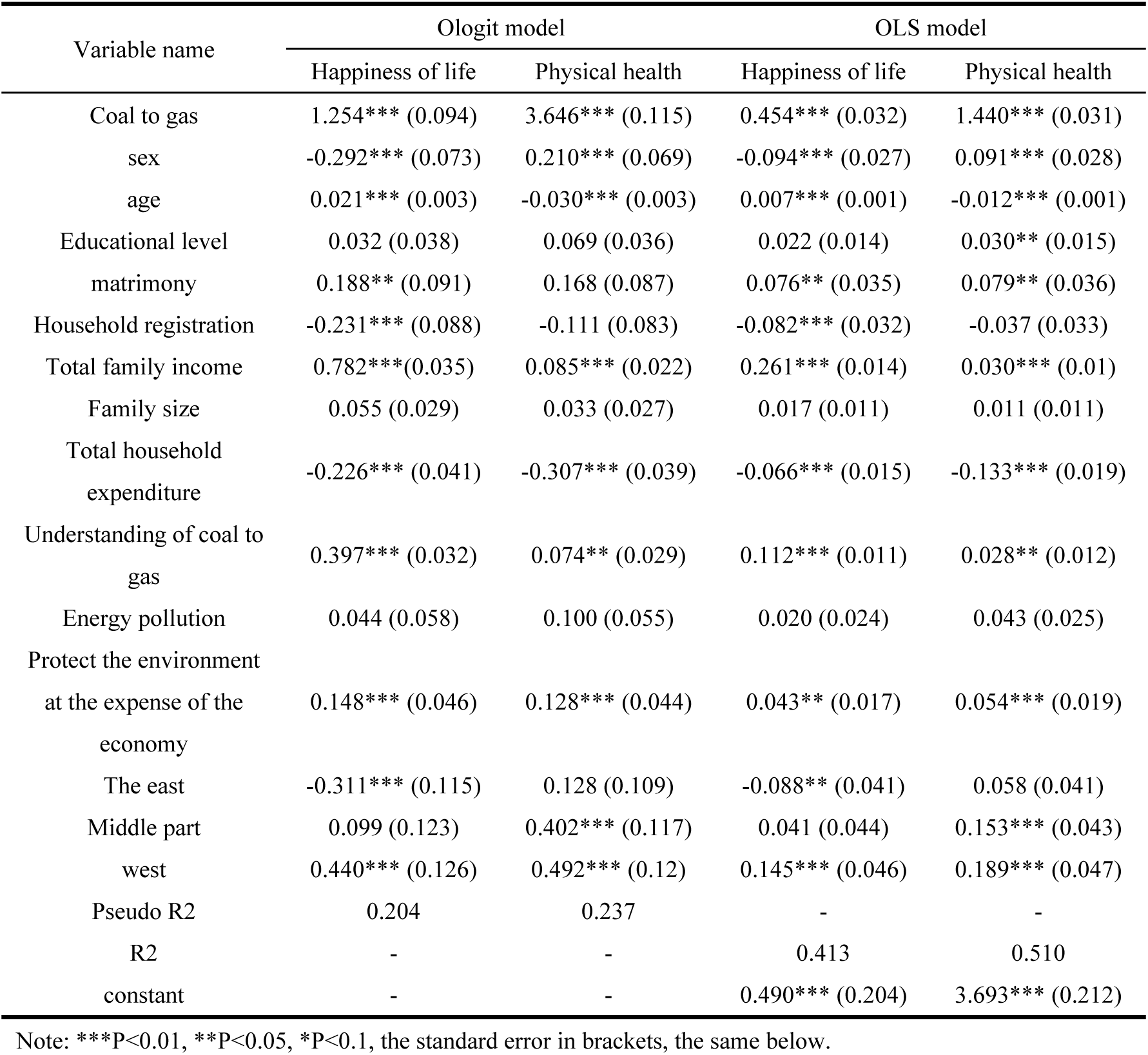
Impacts of coal-to-gas policy on residents’ subjective well-being.

In terms of control variables, gender has a significant negative impact on residents’ happiness at 1% level, and a positive impact on residents’ physical health at 1% level, indicating that men’s happiness is generally lower than women’s, which may be related to changes in life cycle happiness and social expectations. In terms of health, men seem to be better than women, which may be related to physiological structure and genetic differences, but they may face more challenges in mental health and bad lifestyle habits, which is similar to the conclusion of the study of Nemakek et al. (2019).

The results in Table 4 show that the older the residents are, the happier they are, while the worse their physical condition is, which may be attributed to the positive changes at the psychological level and the changes in their social roles, such as the residents’ greater appreciation of interpersonal relationships, pursuit of interests and social respect, which all enhance their happiness. However, with the increase of age, physical functions gradually decline and chronic diseases increase, leading to a decline in physical condition, which is similar to the conclusion of Zahida et al. (2020).

The marital status of 5% promotes the happiness of residents, indicating that the better the marital status, the higher the happiness of residents. The possible reason is that a stable marital relationship provides emotional support and dependence, role identity enhances the sense of self-realization of individuals, mutual help between spouses improves the quality of life, and similar interests deepen the emotional connection between spouses. These work together to improve residents’ happiness, which is similar to the conclusion of Ma et al. (2024)].

Household registration negatively affects residents’ happiness of life at 1% level, that is, residents with rural household registration are less happy than those with urban household registration. The possible reason is that rural residents’ income and living standard are different from those of urban residents due to the lagging economic development in rural areas, and rural residents cannot enjoy the same level of services as urban residents in education, medical care, social security and other aspects. At the same time, insufficient living environment and facilities in rural areas, such as inconvenient transportation and poor living conditions, also have a negative impact on happiness, which is consistent with the conclusion of Zhang et al. (2024).

Household income at 1% positively promotes residents’ well-being, while household expenditure at 1% negatively significantly affects residents’ well-being. In other words, the higher the household income, the higher the residents’ well-being. The higher the household expenditure, the worse the household well-being, possibly for the following reasons: Higher income generally leads to higher quality of life and economic security, which in turn increases well-being; However, the relationship between family expenditure and family well-being is more complicated. If excessive expenditure is improperly distributed, especially when it is mainly used for non-essential consumption or leads to debt pressure, family well-being may be reduced, which is consistent with the research conclusion of Palash et al. (2024)

The understanding level of coal to gas and the perception of sacrificing economic benefits for environmental protection have a significant positive impact on residents’ happiness and health status at 1% level. The possible reason is that the clearer residents’ understanding of the coal to gas policy and their perception of sacrificing economic benefits for environmental protection, the easier they are to understand the importance of improving air quality and the urgency of environmental protection. They have a higher pursuit of quality of life, which in turn improves their happiness and health. This is consistent with the conclusion of Qian et al. (2022).

In the eastern region, a 1% level has a negative impact on residents’ happiness. The possible reasons are that the more developed the eastern economy, the pressure of fierce competition, especially in the fields of work and education, and the psychological pressure brought about by the high expectations of the society for economic achievement, at the same time, the high cost of living, the decline of environmental quality also exacerbated the economic pressure and the concern about the quality of life, which together reduce the happiness of residents. This is consistent with the findings of Liu and Hu (2021).

In the central region, at the level of 1%, there is a significant positive impact on the physical health of residents. This is mainly due to the in-depth implementation of the new rural cooperative medical care policy, the improvement of health education and literacy, the improvement of life style and the enhancement of economic conditions. The combined effect of these factors has played a positive role in guaranteeing residents’ medical services, enhancing health awareness, improving living habits and improving the quality of life. This is consistent with the findings of Sun et al. (2021).

In the western region, at the level of 1%, there is a significant positive impact on residents’ happiness and physical health. Economic development leads to the steady growth of residents’ income; meanwhile, the enhancement of social integration and ethnic exchanges promotes the cohesion and harmonious atmosphere of the community; the optimal allocation of medical resources enables residents to enjoy higher quality medical services; the popularization of health education significantly enhances residents’ health awareness. In turn, it promotes the improvement of residents’ lifestyle, and these comprehensive factors jointly promote the significant improvement of residents’ well-being in the western region. This is consistent with the results of Liu et al.(2023).

### 4.2 Inverted U-shaped empirical analysis of pipeline availability on residents’ well-being

The regression model in Table 5 provides an in-depth analysis of the specific impact of the availability of natural gas pipelines on residents’ well-being. In the sample, only 978 residents believed that the gas pipeline was usable, accounting for only 31.8%. Among these groups of residents who were able to perceive the availability of a gas pipeline, the distance of the gas pipeline from the place of residence had significantly different effects on well-being. Specifically, when the gas pipeline is supplied to the household "at zero distance", or "supplied to the household but at a relatively close distance (1-3 km)", or even in the extreme case of "more than 10 km", the well-being of residents is significantly positive. However, when the pipeline was "a little further (3-5 km)" or "a little further (5-10 km)", there was a significant decline in residents’ well-being. In addition, with the gradual weakening of the availability of natural gas pipelines, their impact on residents’ well-being first showed a downward trend and then rebounded, forming a "U-shaped" impact. This finding strongly validates hypothesis 2, that is, the impact of the availability of natural gas pipelines on residents’ well-being is closely related to its difficulty and presents a non-linear correlation.

**Table. 5.** U-shaped influence of coal-to-gas supply distance on residents’ happiness.

| Variable name | Zero distance,<br>supply to the<br>household | supply to the household<br>is relatively close, 1-3<br>km | a little far,<br>3-5 km | far, 5-10<br>km | very far, more<br>than 10 km |
| --- | --- | --- | --- | --- | --- |
| Happiness of life | 0.150***<br>(0.055) | 0.069***<br>(0.008) | -0.736***<br>(0.088) | -0.129***<br>(0.014) | 0.505***<br>(0.122) |
| Control variable | Control | Control | Control | Control | Control |
| Local control<br>variable | Control | Control | Control | Control | Control |
| R2 | 0.478 | 0.475 | 0.509 | 0.475 | 0.483 |
| constant | 0.562*<br>(0.361) | 0.525***<br>(0.036) | 1.048***<br>(0.355) | 0.559***<br>(0.162) | 0.520***<br>(0.159) |
| Variable name | Zero distance,<br>supply to the<br>household | supply to the household<br>is relatively close, 1-3<br>km | a little far,<br>3-5 km | far, 5-10<br>km | very far, more<br>than 10 km |
| Physical health | 0.175**<br>(0.072) | 0.015***<br>(0.004) | -0.759***<br>(0.120) | -0.175***<br>(0.018) | 0.590***<br>(0.162) |
| Control variable | Control | Control | Control | Control | Control |
| Local control<br>variable | Control | Control | Control | Control | Control |
| R2 | 0.247 | 0.243 | 0.274 | 0.244 | 0.253 |
| constant | 4.007***<br>(0.477) | 3.981***<br>(0.480) | 4.506***<br>(0.476) | 4.006***<br>(0.479) | 3.958***<br>(0.476) |

### 4.3 Marginal effect test

Table 6 reports the marginal impact of the coal-to-gas policy on residents’ well-being. From the perspective of the marginal effect of coal to gas, the implementation of coal to gas policy has improved the probability of residents’ well-being. Residents’ participation in the coal to gas policy has a significant negative correlation with residents’ "very unhappy", "relatively unhappy" and "general", while it has a significant positive correlation with residents’ "relatively happy" and "very happy", namely: When residents increase the degree of participation in replacing coal with gas by one unit, the subjective well-being and happiness of residents increase by 1.254 units, while the probabilities of "very unhappy", "relatively unhappy" and "average" decrease by 1.5%, 9% and 8.5% respectively, while the probabilities of "relatively happy" and "very happy" increase by 5.1% and 13.7%. Similarly, residents’ participation in the coal to gas policy has a significant negative correlation with residents’ "very unhealthy" and "relatively unhealthy", while it has a significant positive correlation with residents’ "average", "relatively healthy" and "very healthy", namely: When residents increase the degree of participation in replacing coal with gas by one unit, the subjective well-being and happiness of residents increase by 3.646 units, while the probabilities of "very unhealthy" and "relatively unhealthy" decrease by 13.4% and 40.6% respectively, while the probabilities of "average", "relatively healthy" and "very healthy" increase by 7.7%, 14.0% and 31.4%. It can be seen that the participation in coal conversion to gas can significantly improve the well-being of residents, especially the promotion effect on physical health is more intense and obvious (Zhu et al,2024).

**Table. 6.**
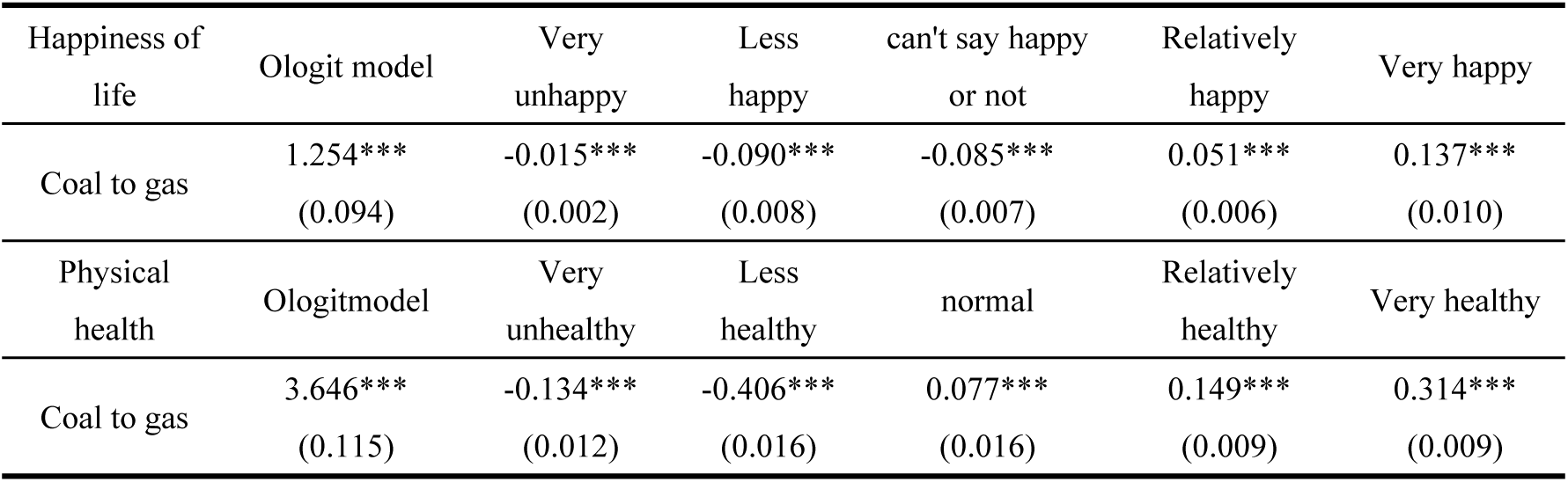
Marginal effects.

### 4.4 Endogeneity test

The OLS regression model is used to deal with the endogenous problem. In terms of the selection of instrumental variables, the participation degree of provincial coal to gas is used as the instrumental variable of the coal to gas policy, and the two-stage estimation of the 2SLS regression model is adopted. The results are shown in Table 7. It can be seen from the table that the instrumental variables of the first stage have a significant positive impact on the core explanatory variable of the coal-gas policy. The F statistic of the first stage is 116.228, which is significantly greater than 10, indicating that there is no weak instrumental variable problem, that is, the selection of instrumental variables is effective. It can be seen from the estimation of the second stage model that the effect of coal to gas on residents’ happiness and physical health is still significantly positive, which is significant at the level of 1%. Therefore, instrumental variables can be explained well in the model.

**Table. 7.** Endogeneity test.

| Variable name | Happiness of life |  | Physical health |  |
| --- | --- | --- | --- | --- |
|  | First stage | Second stage | First stage | Second stage |
| Coal to gas | - | 0.020*** (0.005) | - | 0.914*** (0.127) |
| Instrumental variable | 0.796*** (0.052) | - | 0.796*** (0.052) | - |
| Control variable | Control | Control | Control | Control |
| Local control variable | Control | Control | Control | Control |
| R2 |  | 0.187 |  | 0.461 |
| Wu-Hausman |  | 19.872*** |  | 21.130*** |
| Durbin |  | 19.853*** |  | 21.101*** |
| Wald |  | 690.919*** |  | 1163.818*** |
| Phase one F value |  | 116.228*** |  | 116.228*** |
| Tool variable t value |  | 15.243*** |  | 15.243*** |

### 4.5 Robustness test

1. Propensity score matching method. In order to further verify the robustness of the analysis results, the propensity score matching of the impact of participation in the coal-to-gas policy on residents’ happiness and physical health was analyzed. As shown in Table 8, ATT of samples participating in the coal-to-gas policy and not participating in the coal-to-gas policy after matching was calculated, whether it was nearest neighbor matching, radius matching, nuclear matching and local linear regression matching. Both were significant at the level of 1%. The results of the four matching methods are similar, which reflects the robustness of the matching results to a certain extent. The ATT results all show that, after eliminating the observable systematic differences in samples, participation in the coal-to-gas policy will bring happiness and health effects to residents, further verifying the robustness of the above conclusions.

**Table. 8.** Matching results of PSM propensity score.

| Happiness of life | Treatment group | Control group | ATT | Standard error | T-value |
| --- | --- | --- | --- | --- | --- |
| Nearest neighbor matching | 4.229 | 3.703 | 0.526*** | 0.0339 | 15.53 |
| Radius matching | 4.229 | 3.464 | 0.765*** | 0.029 | 26.08 |
| Nuclear matching | 4.229 | 3.888 | 0.341*** | 0.042 | 8.20 |
| Local linear regression matching | 4.227 | 3.904 | 0.323*** | 0.051 | 6.38 |
| Mean value | - | - | 0.489 | - | - |
| Physical health | Treatment group | Control group | ATT | Standard error | T-value |
| Nearest neighbor matching | 4.309 | 2.841 | 1.467*** | 0.032 | 46.08 |
| Radius matching | 4.309 | 2.762 | 1.546*** | 0.028 | 55.43 |
| Nuclear matching | 4.309 | 2.779 | 1.530*** | 0.037 | 41.75 |
| Local linear regression matching | 4.308 | 2.817 | 1.491*** | 0.048 | 30.99 |
| Mean value | - | - | 1.509 | - | - |

**Table. 9.** Replacement variables.

| Variable name | Binary Logistic model |  |
| --- | --- | --- |
|  | Whether happy or not | Whether healthy or not |
| Coal to gas | 1.718*** (0.143) | 3.959*** (0.146) |
| Control variable | Control | Control |
| Local control variable | Control | Control |
| R2 | 0.443 | 0.573 |
| constant | -10.431*** (0.788) | 2.398** (0.690) |

**Table. 10.** U-shaped test of supply distance of natural gas pipeline. As can be seen from fig. 6 and fig 7, there are significant differences in the probability density distribution of propensity scores before matching between samples of the two groups participating in the coal-to-gas policy (treatment group) and those not participating in the coal-to-gas policy (control group). After propensity score matching, the propensity score probability distribution of the two groups of samples tends to be significantly consistent. After matching, the two groups of samples showed a large range of overlap in propensity scores, which indicated that the common support conditions were satisfied. The common support condition ensured that the two groups of samples had certain similarity in propensity scores when comparing the treatment group and the control group, thus improving the validity and credibility of the comparison results. The successful application of propensity score matching not only eliminates the systematic bias in the original data, but also improves the comparability between samples. The advantage of this approach is that it can assess the impact of coal-to-gas policies on target groups based on a single dimension, propensity score, without taking into account other potential confounding factors. Therefore, the matched samples provide us with a more accurate and reliable framework to evaluate the actual effects of the coal-to-gas policy in promoting the transformation of energy structure and improving environmental quality.
2. Substitution variable method. The robustness test was carried out using alternative variables, and the data were reorganized by referring to the study of Deng et al. (2023). "Very unhappy, relatively unhappy, not happy" was regarded as "unhappy", "relatively happy, very happy" was regarded as "happy", and the values were assigned as 0 and 1 respectively. Similarly, "very unhealthy, relatively unhealthy, not healthy or unhealthy" is regarded as "unhealthy", and "relatively happy or very happy" is regarded as "healthy", with values of 0 and 1 respectively. Since the variables at this time were 0-1 variables, binary Logistic model and OLS model were used to re-regression, and the results showed that the participation in the coal-to-gas policy had a positive impact on residents’ well-being at the 1% level, verifying the robustness of the above conclusions.
3. Placebo test. Although the robustness test shows that the benchmark regression results in this paper are reliable, it cannot exclude the randomness of the results and the potential influence of other unobservable factors. Based on this, according to the study of Cai et al. (2016), this paper conducted a placebo test using a randomly generated experimental group, repeated 1000 times, obtained the probability of the estimated coefficient of the impact of the coal-to-gas policy on residents’ happiness and physical health based on the false experiment, and then judged the reliability of the research results. The results of placebo test are shown in fig. 8 and fig. 9. The estimated coefficients are distributed near 0, and the P-value is mostly greater than 0.1, while the T-value is concentrated near 0, indicating that the setting of the model is reliable and the previous results are relatively robust.
4. U-shaped test of the supply distance of natural gas pipelines. The U-shaped impact of natural gas pipeline availability on residents’ well-being has been verified above. Referring to the study of Yang et al. (2022), the direct impact of natural gas pipeline supply distance and the square distance of natural gas supply distance on residents’ well-being is empirically analyzed. The results show that, regardless of 0logit model or OLS model, The availability of natural gas pipelines has a significant impact on residents’ well-being. Specifically, the coefficient of the primary term is significantly negative, and the coefficient of the secondary term is significantly positive, which once again proves that the impact of pipeline availability on residents’ well-being is non-linear and presents a "U-shaped" influence.

| Variable name | Ologit model |  | OLS model |  |
| --- | --- | --- | --- | --- |
|  | Happiness of life | Physical health | Happiness of life | Physical health |
| Gas pipeline supply distance | -1.956*** (0.343) | -1.831*** (0.317) | -0.748*** (0.112) | -0.857*** (0.149) |
| Gas pipeline supply distance squared | 0.390*** (0.066) | 0.344*** (0.061) | 0.138*** (0.021) | 0.160*** (0.028) |
| Control variable | Control | Control | Control | Control |
| Local control variable | Control | Control | Control | Control |
| Pseudo R2 | 0.260 | 0.111 | - | - |
| R2 | - | - | 0.497 | 0.268 |
| constant | - | - | 1.513*** (0.386) | 5.088*** (0.513) |
| Observed value | 978 | 978 | 978 | 978 |

### 4.6 Population heterogeneity analysis

When assessing the impact of the coal-to-gas policy on residents’ well-being, although the preliminary conclusion indicates that participation in the policy can help improve residents’ well-being, this conclusion is mainly based on the average effect of the whole sample and fails to fully take into account the heterogeneity among different residents groups. In order to have a more comprehensive and in-depth understanding of the effect of this policy, this paper analyzes the heterogeneity of participation in the coal-to-gas policy from the perspectives of age, gender education level, annual total household consumption, urban and rural areas and regions. The main reasons are as follows: First, age is a key factor affecting individuals’ acceptance of new technologies and policies. There are differences in cognition, learning speed and adaptability among residents of different ages (Smith and Levinson,2014). Younger people are generally more receptive to new technologies and tend to access and share information via the Internet, while older people may need more time and support to adjust to new lifestyles and energy use. Second, gender differences also influence how residents respond to new technologies and policies. Studies have shown that there are gender differences in energy use habits, environmental awareness and information access (Jones and Kammen,2014). Therefore, gender factors play an important role in the promotion of coal to gas policies and the improvement of residents’ well-being. Education level is an important factor affecting individual cognitive ability, information processing ability and decision-making ability. Residents with a higher level of education are generally more likely to understand the significance and benefits of the coal-to-gas policy, and are able to participate more effectively in the coal-to-gas policy and compare different energy options (Hargreaves and Adkins,2015). Total annual household consumption reflects residents’ economic capacity and willingness to pay for new energy. High-consumption households may be more willing to invest in new energy equipment and technologies, thereby benefiting more quickly from the coal-to-gas policy (Apergis and Payne, 2011). In addition, the annual total household consumption also indirectly reflects the living standards and well-being of residents. The differences between urban and rural areas and regions reflect the imbalance of social and economic development. There are differences in energy infrastructure, policy support and residents’ income in different regions, all of which may affect the implementation effect of the coal-to-gas policy and the improvement of residents’ well-being (Lin and Zhu, 2019). Therefore, the results of this part of the estimates are shown in Table 11.

**fig. 6.**
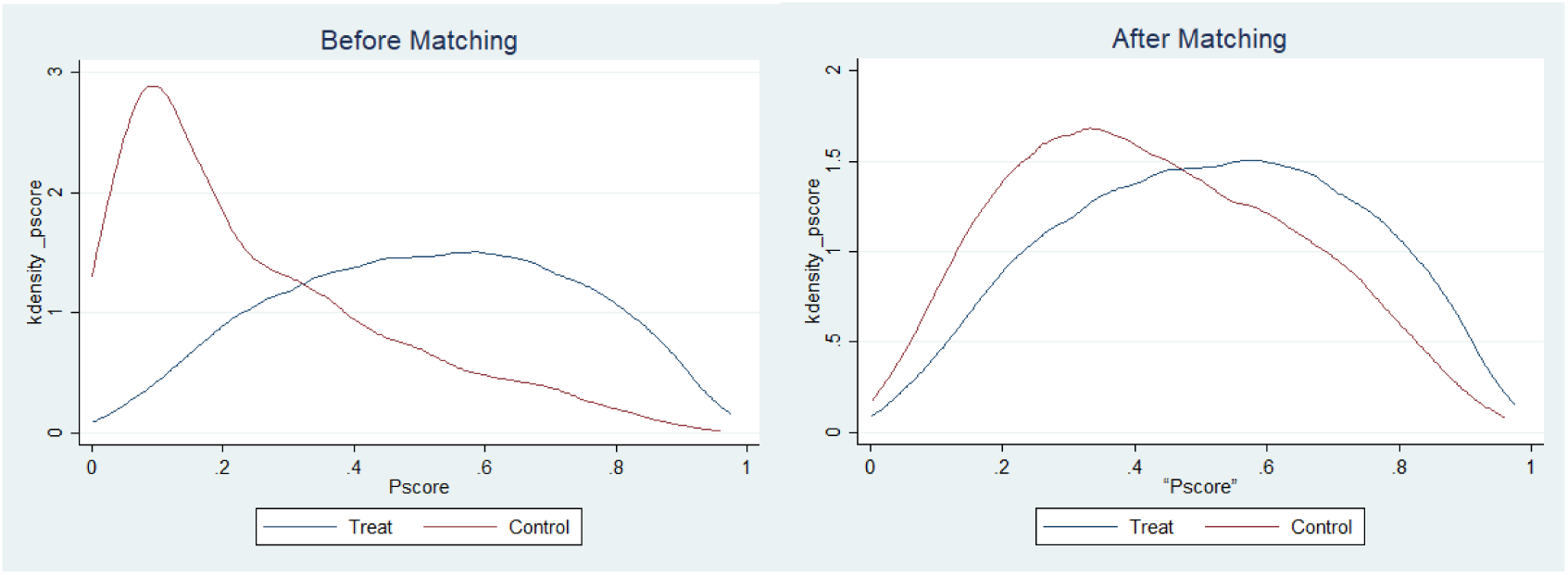
Propensity score of happiness of life matched kernel density map

**fig. 7.**
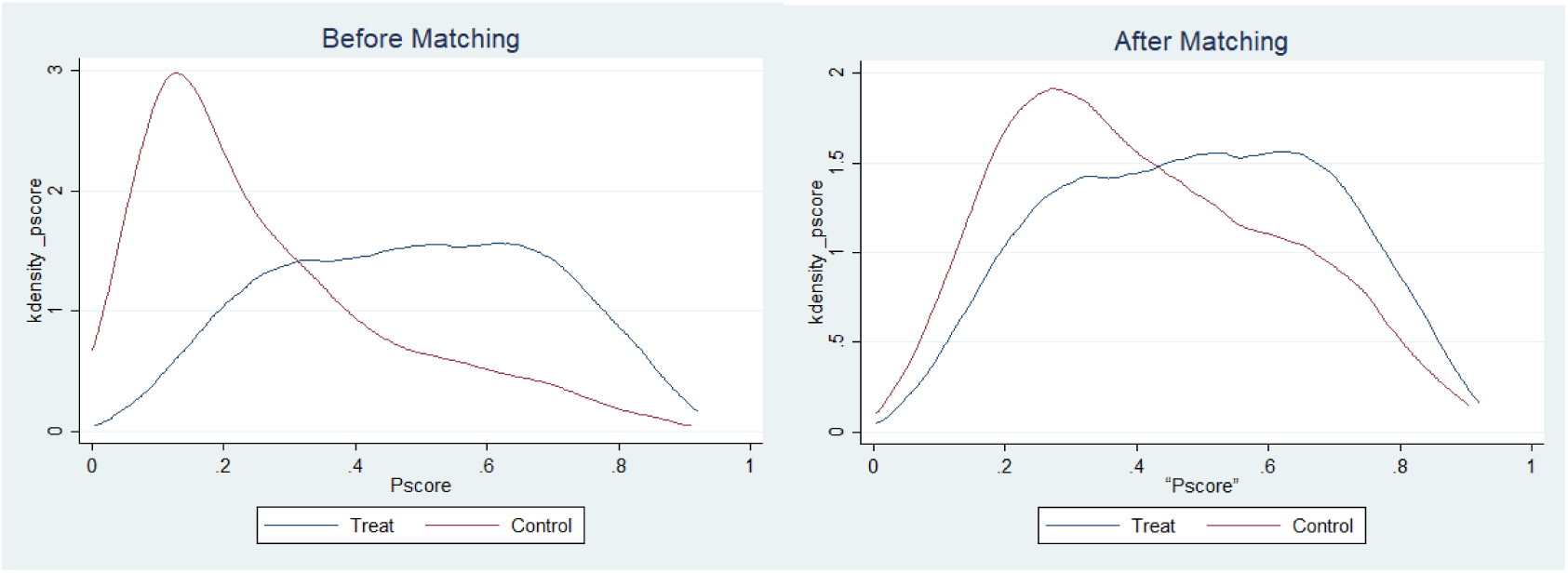
Propensity scores for physical health matched with nuclear density maps

**Fig. 8.**
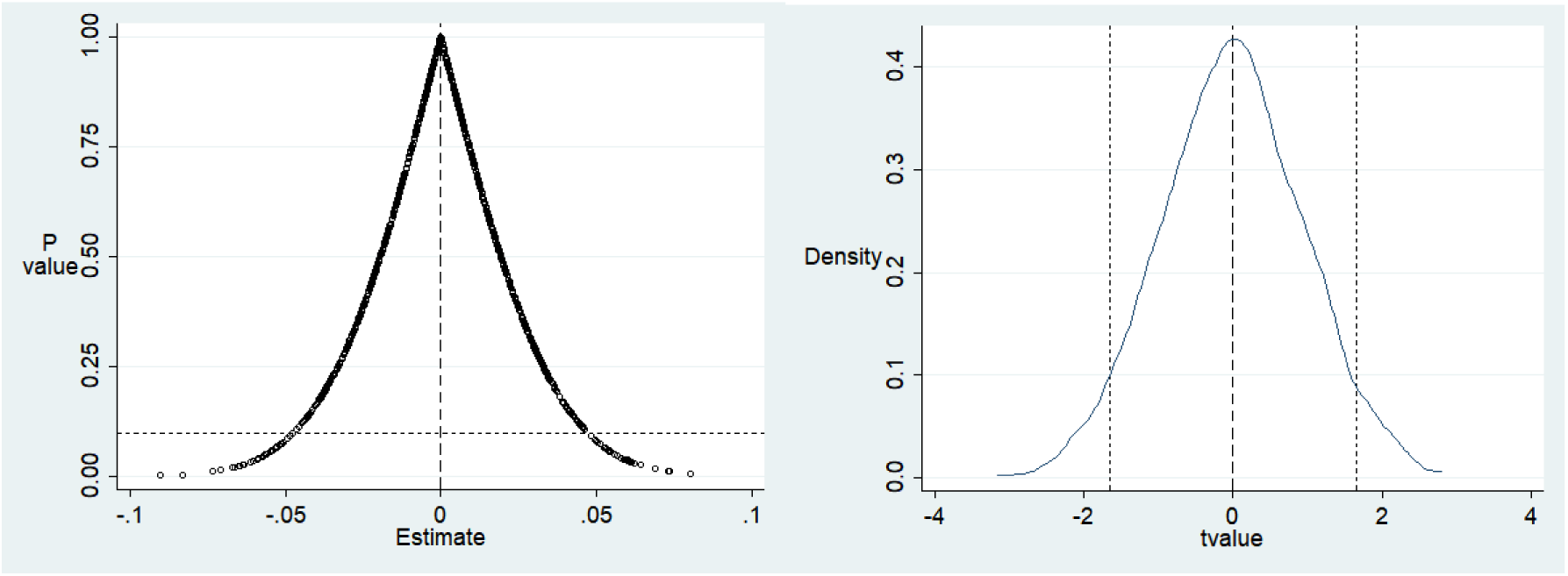
Stabilizer test of the impact of coal-to-gas policy on residents’ happiness of life

**Fig. 9.**
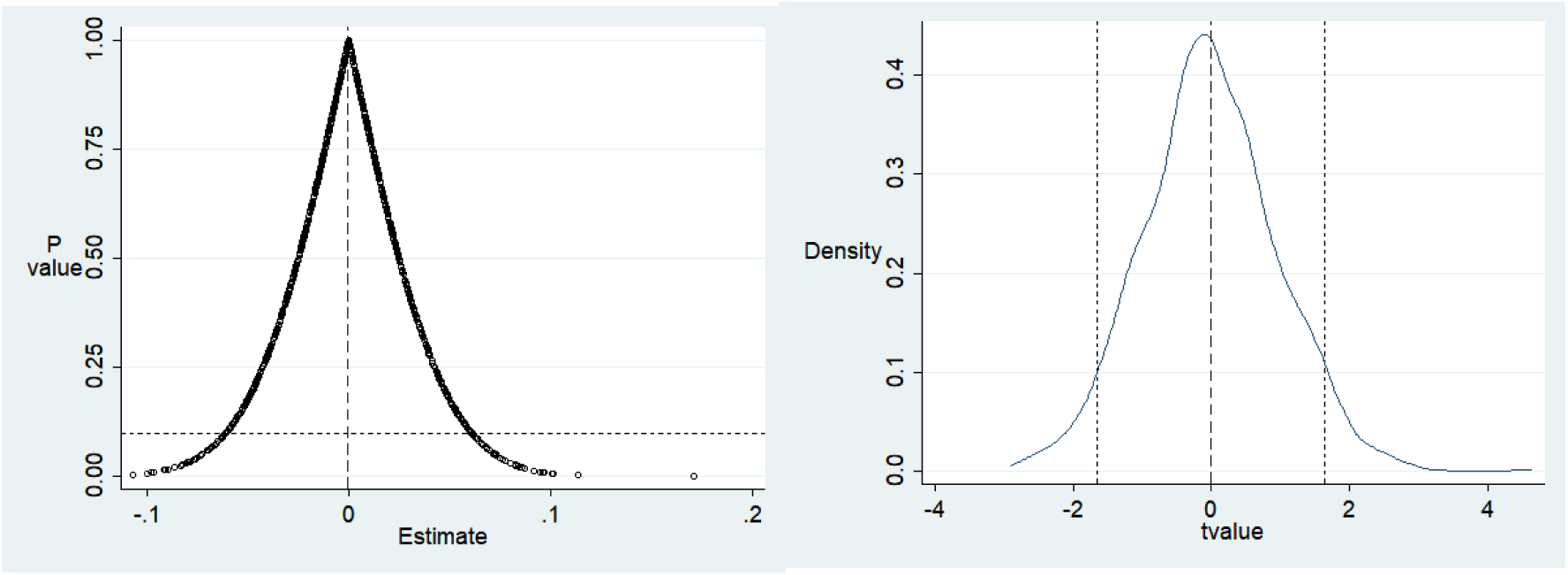
Test of stabilizer involved in the impact of coal-to-gas policy on residents’ physical health

**Table. 11.** Heterogeneity was analyzed by OLS model.

| GroupA | Age > 60 |  | Age ≤ 60 |  |
| --- | --- | --- | --- | --- |
|  | Happiness of life | Physical health | Happiness of life | Physical health |
| coal to gas | 0.474*** (0.070) | 1.397*** (0.063) | 0.585*** (0.041) | 1.527*** (0.040) |
| Control variable | Control | Control | Control | Control |
| Local control variable | Control | Control | Control | Control |
| R2 | 0.225 | 0.416 | 0.284 | 0.491 |
| constant | 2.293*** (0.200) | 2.073*** (0.179) | 2.344*** (0.136) | 2.235*** (0.132) |
| Observed value | 957 | 957 | 2123 | 2123 |
| Group B | Above junior high school |  | Junior high school and below |  |
|  | Happiness of life | Physical health | Happiness of life | Physical health |
| coal to gas | 0.553*** (0.049) | 1.473*** (0.048) | 0.539*** (0.048) | 1.495*** (0.046) |
| Control variable | Control | Control | Control | Control |
| Local control variable | Control | Control | Control | Control |
| R2 | 0.215 | 0.535 | 0.288 | 0.46 |
| constant | 2.876*** (0.208) | 2.711*** (0.2022) | 1.866*** (0.154) | 2.659*** (0.146) |
| Observed value | 1057 | 1057 | 2023 | 2023 |
| Group C | Men |  | Women |  |
|  | Happiness of life | Physical health | Happiness of life | Physical health |
| coal to gas | 0.560*** (0.050) | 1.466*** (0.049) | 0.594*** (0.049) | 1.521*** (0.045) |
| Control variable | Control | Control | Control | Control |
| Local control variable | Control | Control | Control | Control |
| R2 | 0.293 | 0.472 | 0.244 | 0.513 |
| constant | 1.674*** (0.177) | 2.766*** (0.175) | 2.360*** (0.184) | 2.503*** (0.170) |
| Observed value | 1446 | 1446 | 1634 | 1634 |
| Group D | Family high consumption group |  | Family low consumption group |  |
|  | Happiness of life | Physical health | Happiness of life | Physical health |
| coal to gas | 0.512*** (0.057) | 1.505*** (0.057) | 0.600*** (0.045) | 1.490*** (0.041) |
| Control variable | Control | Control | Control | Control |
| Local control variable | Control | Control | Control | Control |
| R2 | 0.282 | 0.534 | 0.246 | 0.477 |
| constant | 2.325*** (0.218) | 1.754*** (0.217) | 1.715*** (0.154) | 2.705*** (0.142) |
| Observed value | 982 | 982 | 2098 | 2098 |
| Group E | village |  | city |  |
|  | Happiness of life | Physical health | Happiness of life | Physical health |
| coal to gas | 0.501*** (0.046) | 1.537*** (0.046) | 0.632*** (0.054) | 1.423*** (0.047) |
| Control variable | Control | Control | Control | Control |
| Local control variable | Control | Control | Control | Control |
| R2 | 0.28 | 0.488 | 0.247 | 0.511 |
| constant | 1.966*** (0.161) | 2.415*** (0.161) | 1.842*** (0.220) | 2.841*** (0.191) |
| Observed value | 1915 | 1915 | 1165 | 1165 |
| Group F | Southern region |  | Northern region |  |
|  | Happiness of life | Physical health | Happiness of life | Physical health |
| coal to gas | 0.501*** (0.057) | 1.416*** (0.056) | 0.699*** (0.047) | 1.601*** (0.041) |
| Control variable | Control | Control | Control | Control |
| Local control variable | Control | Control | Control | Control |
| R2 | 0.247 | 0.383 | 0.302 | 0.610 |
| constant | 2.168*** (0.169) | 2.835*** (0.168) | 1.941*** (0.186) | 2.247*** (0.164) |
| Observed value | 1689 | 1689 | 1391 | 1391 |

In terms of age group, we investigate the impact of coal to gas on the elderly population and non-elderly population. In this paper, the internationally recognized age of 60 is taken as the standard for dividing the elderly population, and the population under 60 is taken as the "young group of farmers" and the population over 60 is taken as the "old group of farmers", that is, the age of 60 is taken as the dividing point of heterogeneity. The results show that participation in the coal to gas policy has a greater impact on the happiness and health status of the non-elderly population than that of the elderly population. It can be seen that the coal to gas policy has a significant impact on the age heterogeneity of the population. The possible reason is that the non-elderly population has a more direct experience of clean and efficient energy use due to their more frequent participation in social activities. Thus improving their quality of life and happiness. However, due to the limitations of living habits and physical conditions, the elderly population has a weak perception of the changes in life brought about by the coal-to-gas policy. Non-elderly people are more likely to accept and adapt to new lifestyles, such as using natural gas instead of coal, which not only reduces environmental pollution, but also improves the convenience and comfort of life, which is consistent with Cosco et al. (2017).

From the perspective of education level, participation in the coal to gas policy has a more significant impact on the happiness of the group above junior middle school and on the Physical health of the group below junior middle school. The possible reasons are as follows: due to the improvement of education background and environmental awareness, the group above junior middle school is more likely to feel the improvement of happiness from the policy; On the other hand, those in junior high school and below are more concerned about the direct impact of policies on daily life and Physical health, which is consistent with Chakraborty et al. (2019).

From the perspective of gender groups, compared with male residents, the change from coal to gas has a more significant impact on the happiness and Physical health of female residents, that is, the happiness of women participating in the change from coal to gas policy will be significantly improved, which is mainly attributed to women’s role positioning in the family, health awareness and participation in housework. Replacing coal with gas can improve household environmental quality, reduce air pollution and reduce health risks, thus significantly improving the happiness and Physical health level of female residents, which is consistent with the research conclusion of Smith et al. (2023).

In terms of the household consumption group, we choose the total household consumption as the grouping criterion: that is, lower than the average total consumption is a low consumption group, and higher than the average total consumption level is a high consumption group. The results show that the participation in the coal to gas policy has a more significant impact on the happiness of the low consumption group and the Physical health of the high consumption group. The possible reasons are as follows: For the low consumption families, the reduction of economic cost and the improvement of environmental quality brought by the policy significantly enhance their happiness; For high-consumption families, policies are more conducive to their pursuit of high-quality life and guarantee of Physical health, which is consistent with the research conclusion of Li et al. (2022).

From the perspective of urban and rural heterogeneity, participation in the coal to gas policy has a more significant impact on the happiness of urban residents and the Physical health of rural residents. The possible reasons are as follows: Due to their higher education level and life quality requirements, urban residents have a deeper understanding and cognition of the environmental protection and life quality improvement brought by the policy, thus increasing their happiness; Since the living environment of rural residents is closely related to the use of coal, the implementation of policies can directly improve their living environment and reduce health risks, which has a more significant impact on Physical health, which is consistent with the conclusion of Wang et al. (2023).

From the perspective of the difference between the south and the north, compared with the southern residents, the change from coal to gas has a more significant impact on the happiness and Physical health of the northern residents, that is, the happiness and Physical health of the northern residents participating in the coal to gas policy will be significantly improved. The possible reasons are as follows: Compared with the southern region, the climate characteristics and social cognition of the northern region make the impact of the coal-to-gas policy more significant. The implementation of the coal-to-gas policy not only improves their heating conditions and living environment, but also significantly improves their happiness and Physical health, which is consistent with the research conclusion of Zhang et al. (2022).

### 4.7 Analysis of the effect of new energy transformation

According to theoretical analysis, the policies involved in the transformation of coal into gas affect residents’ well-being through income substitution effect mechanism, gas supply effect mechanism, gas capital-intensive industry agglomeration effect mechanism, industrial transfer effect mechanism, industrial transfer effect mechanism and environmental effect mechanism. Therefore, this part seeks for appropriate proxy variables and verifies the correctness of the theories, as shown in Table 12-17.

**Table. 12.** Income substitution effect.

| Variable name | Ologit model |  | OLS model |  |
| --- | --- | --- | --- | --- |
|  | Happiness of life | Physical health | Happiness of life | Physical health |
| Coal to gas | 1.414***<br>(0.091) | 3.665***<br>(0.113) | 0.528***<br>(0.031) | 1.532*** (0.03) |
| Total family income | 0.643***<br>(0.029) | 0.071***<br>(0.023) | 0.244***<br>(0.012) | 0.026*** (0.01) |
| Regional gross domestic product | -0.173***<br>(0.06) | 0.164***<br>(0.058) | 0.063***<br>(0.024) | 0.079***<br>(0.026) |
| Coal to gas x total household income | -0.101 (0.063) | -0.076 (0.058) | 0.056 (0.046) | -0.031 (0.022) |
| Coal to gas × regional GDP per capita | 0.011***<br>(0.003) | 0.158***<br>(0.021) | 0.013** (0.05) | 0.103** (0.047) |
| Control variable | Control | Control | Control | Control |
| Local control variable | Control | Control | Control | Control |
| Pseudo R2 | 0.158 | 0.204 | - | - |
| R2 | - | - | 0.360 | 0.460 |
| constant | - | - | 1.634***<br>(0.271) | 1.688***<br>(0.277) |
| Observed value | 3080 | 3080 | 3080 | 3080 |

**Table. 13.** Gas supply effect.

| Variable name | Ologit model |  | OLS model |  |
| --- | --- | --- | --- | --- |
|  | Happiness of life | Physical health | Happiness of life | Physical health |
| Coal to gas | 1.798*** (0.084) | 3.787*** (0.11) | 0.779*** (0.029) | 1.573*** (0.028) |
| City gas supply | 0.122*** (0.037) | 0.194*** (0.037) | 0.054*** (0.016) | 0.074*** (0.015) |
| Coal to gas × city gas supply | 0.039** (0.082) | 0.329** (0.082) | 0.020** (0.009) | 0.110*** (0.031) |
| Control variable | Control | Control | Control | Control |
| Local control variable | Control | Control | Control | Control |
| Pseudo R2 | 0.067 | 0.205 | - | - |
| R2 | - | - | 0.149 | 0.459 |
| constant | - | - | 4.154*** (0.201) | 3.705*** (0.196) |
| Observed value | 3080 | 3080 | 3080 | 3080 |

**Table. 14.** Effect of gas capital-intensive industry agglomeration.

| Variable name | Ologit model |  | OLS model |  |
| --- | --- | --- | --- | --- |
|  | Happiness of life | Physical health | Happiness of life | Physical health |
| Coal to gas | 1.755***<br>(21.043) | 3.769***<br>(0.109) | 0.764***<br>(0.029) | 1.562***<br>(0.027) |
| Production and supply of air, heat, gas and water | -0.021 (0.05) | 0.311***<br>(0.05) | -0.006 (0.025) | 0.131***<br>(0.021) |
| Coal to gas × Air, heat, gas and water production and supply industry | 0.012***<br>(0.003) | 0.384***<br>(0.105) | 0.007***<br>(0.004) | 0.092**<br>(0.039) |
| Control variable | Control | Control | Control | Control |
| Local control variable | Control | Control | Control | Control |
| Pseudo R2 | 0.066 | 0.207 | - | - |
| R2 | - | - | 0.146 | 0.462 |
| constant | - | - | 3.503***<br>(0.168) | 1.876***<br>(0.141) |
| Observed value | 3080 | 3080 | 3080 | 3080 |

**Table. 15.** Effect of industrial transfer.

| Variable name | Ologit model |  | OLS model |  |
| --- | --- | --- | --- | --- |
|  | Happiness of life | Physical health | Happiness of life | Physical health |
| Coal to gas | 1.769***<br>(0.084) | 3.896***<br>(0.112) | 0.769***<br>(0.03) | 1.589***<br>(0.027) |
| Primary industry | 0.023 (0.043) | 0.307***<br>(0.043) | 0.009 (0.02) | 0.121***<br>(0.017) |
| Secondary industry | 0.031 (0.083) | -0.261***<br>(0.083) | 0.002 (0.038) | -0.101***<br>(0.035) |
| Tertiary industry | -0.089 (0.086) | 0.307***<br>(0.085) | -0.027 (0.038) | 0.133***<br>(0.035) |
| Residents participate in coal to gas × primary industry | 0.002**<br>(0.001) | 0.465***<br>(0.093) | 0.008**<br>(0.004) | 0.125***<br>(0.035) |
| Residents participate in coal to gas × secondary industry | -0.220***<br>(0.080) | -0.245***<br>(0.86) | -0.132***<br>(0.051) | -0.114**<br>(0.049) |
| Residents participate in coal to gas × tertiary industry | -0.222**<br>(0.094) | -0.543**<br>(0.183) | -0.126**<br>(0.053) | -0.247**<br>(0.069) |
| Control variable | Control | Control | Control | Control |
| Local control variable | Control | Control | Control | Control |
| Pseudo R2 | 0.066 | 0.213 | - | - |
| R2 | - | - | 0.147 | 0.471 |
| constant | - | - | 3.697***<br>(0.321) | 0.905***<br>(0.27) |
| Observed value | 3080 | 3080 | 3080 | 3080 |

**Table. 16.** Effects of urbanization.

| Variable name | Ologit model |  | OLS model |  |
| --- | --- | --- | --- | --- |
|  | Happiness of life | Physical health | Happiness of life | Physical health |
| Coal to gas | 1.770***<br>(0.084) | 3.885***<br>(0.112) | 0.770***<br>(0.03) | 1.588***<br>(0.027) |
| urbanization | -0.196 (0.18) | -1.388***<br>(0.181) | -0.086 (0.083) | 0.534***<br>(0.073) |
| Residents participate in coal to gas ×<br>urbanization | 0.044**<br>(0.020) | 2.255***<br>(0.383) | 0.022* (0.011) | 0.637***<br>(0.143) |
| Control variable | Control | Control | Control | Control |
| Local control variable | Control | Control | Control | Control |
| Pseudo R2 | 0.066 | 0.211 | - | - |
| R2 | - | - | 0.146 | 0.466 |
| constant | - | - | 3.815***<br>(0.341) | 4.955***<br>(0.302) |
| Observed value | 3080 | 3080 | 3080 | 3080 |

**Table. 17.**
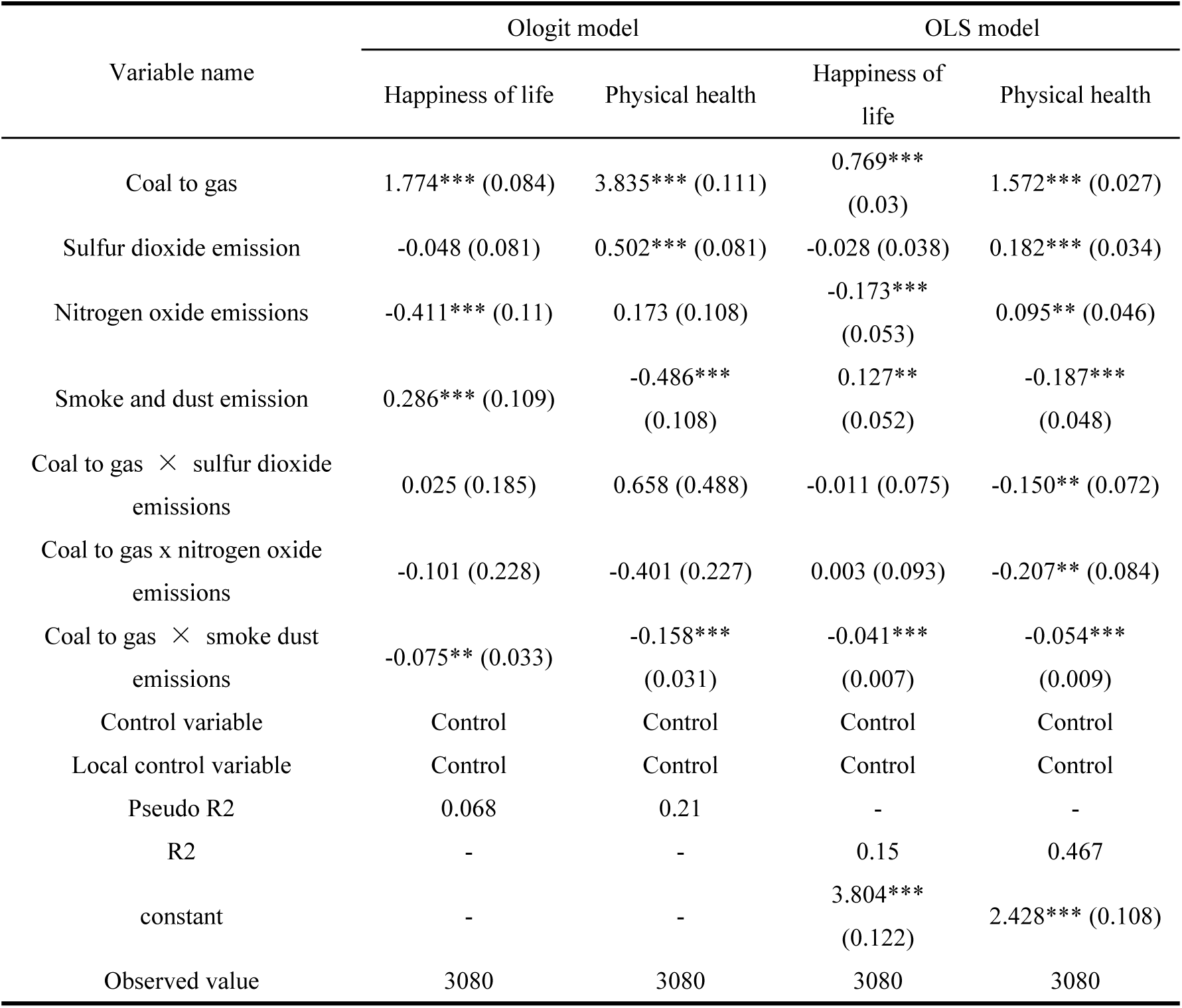
Environmental effects.

| Variable name | Ologit model |  | OLS model |  |
| --- | --- | --- | --- | --- |
|  | Happiness of life | Physical health | Happiness of life | Physical health |
| Coal to gas | 1.774*** (0.084) | 3.835*** (0.111) | 0.769***<br>(0.03) | 1.572*** (0.027) |
| Sulfur dioxide emission | -0.048 (0.081) | 0.502*** (0.081) | -0.028 (0.038) | 0.182*** (0.034) |
| Nitrogen oxide emissions | -0.411*** (0.11) | 0.173 (0.108) | -0.173***<br>(0.053) | 0.095** (0.046) |
| Smoke and dust emission | 0.286*** (0.109) | -0.486***<br>(0.108) | 0.127**<br>(0.052) | -0.187***<br>(0.048) |
| Coal to gas × sulfur dioxide<br>emissions | 0.025 (0.185) | 0.658 (0.488) | -0.011 (0.075) | -0.150** (0.072) |
| Coal to gas x nitrogen oxide<br>emissions | -0.101 (0.228) | -0.401 (0.227) | 0.003 (0.093) | -0.207** (0.084) |
| Coal to gas × smoke dust<br>emissions | -0.075** (0.033) | -0.158***<br>(0.031) | -0.041***<br>(0.007) | -0.054***<br>(0.009) |
| Control variable | Control | Control | Control | Control |
| Local control variable | Control | Control | Control | Control |
| Pseudo R2 | 0.068 | 0.21 | - | - |
| R2 | - | - | 0.15 | 0.467 |
| constant | - | - | 3.804***<br>(0.122) | 2.428*** (0.108) |
| Observed value | 3080 | 3080 | 3080 | 3080 |

We tested the potential mediators of coal to gas and happiness and Physical health. The results show the interaction between coal to gas and the regulatory factors in Table 12. The interaction item between coal to gas and regional per capita GDP has a significant impact at the level of 1%, indicating that the improvement of regional per capita GDP can effectively improve residents’ happiness and Physical health, that is, coal to gas has a significant income substitution effect, which verifies the correctness of hypothesis 3.

It can be seen from Table 13 that the interaction terms between coal to gas and urban gas supply have a significant influence at the 5% level, indicating that the higher the urban gas supply, the higher the happiness and health degree of residents participating in coal to gas, which proves that the energy supply effect of coal to gas is relatively significant, and verifies the correctness of hypothesis 4.

It can be seen from Table 14 that the interaction terms between coal to gas and regional air power consumption have a significant influence at the 1% level, indicating that the higher the regional air power consumption, the higher the happiness and Physical health of residents participating in the coal to gas transformation, indicating that the consumption effect of coal to gas transformation is more significant, verifying the correctness of hypothesis 5.

As can be seen from Table 15, the interaction between the transformation from coal to gas and the number of legal persons in the primary industry is significantly positive, indicating that the transformation from coal to gas can significantly improve the happiness and Physical health of the primary industry group. The interaction between the transformation from coal to gas and the number of legal persons in the secondary industry is significantly negative, and the interaction between the transformation from coal to gas and the number of legal persons in the tertiary industry is significantly negative. It indicates that the transformation from coal to gas significantly reduces the well-being and Physical health of the groups in the secondary and tertiary industries, indicating that the transformation from coal to gas has brought about a significant industrial structure transfer effect, and China’s industrial structure is shifting from the primary industry to the tertiary industry, which verifies the correctness of hypothesis 6.

As can be seen from Table 16, the interaction term between the transformation from coal to gas and the proportion of urban population at the end of 2017 is significantly positive, indicating that the improvement of urbanization level effectively improves the positive impact of the transformation from coal to gas on residents’ happiness and Physical health, indicating that the transformation from coal to gas has a significant urbanization effect, and verifying the correctness of hypothesis 7.

As can be seen from Table 17, the interaction term between coal to gas and smoke and dust is significantly negative at the 5% level, indicating that coal to gas can improve residents’ happiness and Physical health by reducing the emission of smoke and dust in the air, thereby improving the environment, indicating that coal to gas has a significant environmental improvement effect, and verifying the correctness of hypothesis 8.

## 5 Conclusions and policy recommendations

Based on CGSS data of 2018, this paper studies the impact of participation in the coal-to-gas policy on residents’ subjective well-being and physical health, marginal effect, endogeneity, robustness, group heterogeneity and the effect of new energy transformation, and analyzes the impact of the availability of natural gas pipelines, a key variable, on residents’ well-being. The main conclusions of this study can be summarized as follows: (1) Through the application of ordered logistic regression (Ologit) model and least squares (OLS) model, it was verified that the coal-to-gas policy had a significant and positive promoting effect on residents’ subjective well-being and physical health, and passed the endogeneity test. Multiple verification methods such as propensity score matching (PSM), replacement variable method and Robustness Checks are used to ensure the robustness and reliability of the conclusions obtained. The results of marginal effect analysis show that the promotion effect of the participation degree of coal to gas policy on physical health is more intense and obvious. (2) Only 31.8% of residents believe that natural gas pipeline is available, and then the impact of availability of natural gas pipeline on residents’ well-being is analyzed, revealing its significant nonlinear relationship, which is specifically manifested as a "U" -shaped feature. Moderate improvement or significant expansion of natural gas pipeline coverage can have a positive impact on residents’ well-being to some extent. (3) The impact of the coal-to-gas policy on residents’ well-being has significant group heterogeneity. Specifically, among young people, women and residents of northern regions, the policy has had a stronger positive welfare effect. In particular, for people with lower education, higher consumption power and living in rural areas, the coal-to-gas policy has significantly improved their health. However, for the groups with high education, high consumption and urban residents, their subjective well-being has been significantly improved. (4) The positive impact of the coal to gas policy is diverse and far-reaching. It not only injects new vitality into regional economic development by increasing regional per capita GDP, increasing gas supply, promoting the agglomeration of capital-intensive industries and promoting the transformation of industrial structure from primary industry to tertiary industry; At the same time, the policy has also significantly reduced the emissions of smoke and dust in the air, making a positive contribution to improving the quality of the ecological environment, and thus improving the overall well-being of residents.

Based on the above research content, this paper puts forward the following policy recommendations: First, the government should continue to promote the coal-to-gas policy, including strengthening policy publicity, improving residents’ understanding and acceptance of the coal-to-gas policy, increasing financial investment, and ensuring the smooth progress of the coal-to-gas project, especially in the areas that have not been popularized or have low coverage, priority should be given to the construction of the coal-to-gas project. Establish a sound policy monitoring and evaluation system, regularly check and evaluate the implementation of policies, timely identify problems and take measures to solve them, but also strengthen cooperation with relevant departments, form policy forces, and jointly promote the in-depth implementation of the coal to gas policy. In addition, the government should also encourage residents to actively participate in the formulation and implementation of policies to improve the transparency and fairness of policies. Secondly, strengthen the optimization of the layout of natural gas pipelines. When planning natural gas pipelines, it is necessary to consider both the length and coverage of the pipelines, as well as the density and distribution of the pipelines, so as to ensure the moderate improvement of pipeline coverage to maximize its positive impact on the well-being of residents. It is also necessary to strengthen the maintenance and maintenance of pipeline facilities to ensure the safe and stable operation of the pipelines. Clearly optimize the layout of natural gas pipelines, make up for weaknesses, give play to the resource advantages of the western region, strengthen pipeline construction, improve the pipeline network in the eastern region, enhance supply capacity, and promote coordinated development among regions; Thirdly, we should pay special attention to the heterogeneity of groups and formulate differentiated policies and strategies according to the characteristics of different groups. For rural areas, groups with low education and high consumer groups, the government should increase the publicity of the coal-to-gas policy, provide necessary subsidies and support, and encourage them to actively participate in the coal-to-gas project. For urban residents and groups with high education, the government should pay attention to the improvement of their subjective well-being, and meet their needs by improving the living environment and improving the quality of public services. Pay more attention to special groups, such as the elderly and the disabled, to ensure that they can equally enjoy the benefits of the coal to gas policy; Finally, promote economic restructuring and industrial upgrading, and strengthen ecological and environmental protection. By providing tax incentives, land support and other policy measures, attract more capital-intensive industries to settle down, promote the transformation of industrial structure from the primary industry to the tertiary industry, encourage the development of service industries and high-tech industries, and inject new vitality into regional economic development. We will step up efforts to protect the ecological environment, strengthen supervision and treatment of polluting enterprises, ensure that the environmental benefits of the coal-to-gas policy are fully utilized, encourage residents to use renewable energy sources such as solar and wind energy, further reduce pollutant emissions, and improve the quality of the ecological environment.

## 6 Discussion

In the process of in-depth analysis of the impact of coal-to-gas policies on residents’ well-being, this paper greatly enriches the theoretical framework in the field of policy evaluation. Through the construction of a multi-dimensional analysis framework, the complex impact of the coal-to-gas policy is discussed comprehensively and carefully. In particular, this paper reveals for the first time the subtle nonlinear relationship between the availability of natural gas pipelines and residents’ well-being, deepening our understanding of the complexity of policy effects and providing policy makers with more refined regulatory strategies. At the same time, we deepen the understanding of the heterogeneity of different groups in the policy response, and examine the influence of age, gender, region and other factors on the policy effect. In terms of the theory of new energy transformation, the evaluation of the positive role of the coal-to-gas policy in promoting the transformation of energy structure, promoting economic restructuring and industrial upgrading, and the contribution of the policy in ecological environmental protection, provides a new perspective for the research and practice of new energy policy, and provides a strong theoretical support and practical guidance for realizing the sustainable development of society. In the future, the long-term social and economic benefits of the coal-to-gas policy and its dynamic changes can be deeply studied, the impact of the policy on residents’ well-being and economic structure can be evaluated through long-term tracking and analysis, and the continuity and trend of the policy effect can be analyzed, and the dynamic adjustment of the policy and its impact can be explored in combination with the changes of the social and economic environment.

## Data Availability

0

0

## Acknowledgments

This work was supported by National Natural Science Foundation of China (72263018).Science and Technology Innovation Project of Chinese Academy of Agricultural Sciences (10-IAED-RC-01-2024). Jiangxi Social Sciences Fund Project (22SH16).Science and technology research project of Jiangxi Provincial Department of Education (GJJ2200450).

## Notes

### Competing Interest Statement

The authors have declared that no competing interests exist.

### Clinical Trial

0

### Clinical Protocols

0

### Funding Statement

Yes

### Author Declarations

0

